# Study of the associations between short telomeres, sex hormones and pulmonary fibrosis

**DOI:** 10.1101/2022.09.29.22280270

**Authors:** Anna Duckworth, Katherine S. Ruth, Julia K. Prague, Anne-Marie Russell, Howard Almond, John Conway, Robin N Beaumont, Andrew R Wood, Susan Martin, Katie Lunnon, Mark A. Lindsay, Anna Murray, Michael A. Gibbons, Jess Tyrrell, Chris J. Scotton

## Abstract

**Background:** Pulmonary fibrosis (PF) is an incurable fibrotic lung disease with limited treatment options and a high mortality. Evidence is growing that short telomeres cause both heritable and idiopathic pulmonary fibrosis (IPF). Based on survival data, we hypothesised that sex hormones are protective against premature telomere attrition and could influence PF disease onset and/or progression.

**Methods:** Associations between IPF, sex hormone concentrations and measured leukocyte telomere length (LTL) were examined for unrelated UK Biobank participants of European ancestry with a diagnosis of IPF (415 females, 718 males) against controls (204,321 females, 174,254 males). Polygenic risk scores were used to explore causality between sex hormone indices, LTL and disease.

**Findings:** Strong associations were found between IPF and LTL. For females, higher odds of having IPF was associated with early menopause and premature ovarian failure. Menopause age correlated positively with both age of IPF diagnosis and age of death. For males, IPF prevalence and stages of disease were associated with serum bioavailable testosterone concentrations. For both sexes, evidence of lower concentrations of sex hormones was associated with shorter LTL. Genetic analysis also inferred bi-directional causal links between sex hormone binding globulin concentration, which impacts free testosterone concentration, and LTL in males.

**Interpretation:** Our findings suggest that higher sex hormone concentrations protect against IPF onset and progression, possibly by slowing telomere shortening. Hormonal supplementation may delay or prevent disease onset for those with telomere-associated PF risk and improve disease prognosis. This warrants further exploration in a randomised controlled trial.

**Funding:** Medical Research Council.

## Introduction

Idiopathic pulmonary fibrosis (IPF) is a complex and incurable fibrotic lung disease with increasing prevalence and mortality in the UK. Currently IPF accounts for around 5300 UK deaths each year^1^ (62% male^2^) and occurs later in life when sex hormone concentrations are lower.

Circulating sex steroid concentrations are regulated by the hypothalamic-pituitary-gonadal axis in a system of negative feedback to maintain equilibrium throughout life. In men, 1.2-2.9% of total circulating testosterone (depending on age) is freely available to activate testosterone-responsive cells, 33-54% is bound with low affinity to serum albumin (which together constitute ‘bioavailable’ testosterone^3^) and the remainder is tightly bound to sex hormone binding globulin (SHBG)^4^. Similar control of oestrogen bioavailability is seen in women, where the majority is bound to albumin and SHBG. SHBG concentrations vary between, and within, individuals.

Since 2007, evidence has grown detailing the association of pulmonary fibrosis (PF) with short telomeres, the DNA sequences that protect the ends of chromosomes from aberrant repair and inter-chromosomal fusion^5,6^. Familial clustering of PF in patients with dyskeratosis congenita (DC), a rare heritable bone marrow failure syndrome, first implicated mutations in the telomere maintenance genes including *TERT* (telomerase reverse transcriptase) and *TERC* (telomerase RNA component)^5^. Telomerase, which catalyses telomere elongation^7^, is expressed in stem cells, including lung alveolar epithelial cells, where its insufficiency precipitates a cascade of replicative senescence, aberrant wound healing and tissue fibrosis. Recent evidence suggests that short telomeres also contribute to idiopathic PF risk^8^ and associate with worse survival^9^. Using Mendelian randomisation (MR), we recently inferred that telomere length is causally linked with IPF (OR=5.81, 95%CI:3.56-9.50, P=2.19×10^−12^)^10^.

DC and congenital aplastic anaemia (also frequently associated with *TERT* and *TERC* mutations) represent severe telomere biology disorders (TBDs) and both have been treated effectively with androgens since the 1960s ^11,12,13^, although case reports are mixed^14^ and concern exists regarding increased risk of malignancy due to the association between upregulated telomerase and tumourigenesis^15^. However, a recent review of fibrosis in the kidney, liver, lung and heart concluded that sex-related differences in the pathogenesis strongly suggest a role for sex hormones in modulating disease progression^16^. Evidence from androgen deprivation therapy in prostate cancer suggests that the detrimental impact of reduced sex hormone levels is restricted to those at risk for TBDs^17^.

In general, women have longer telomeres^18^, lower PF prevalence^19^ and slower PF progression than men^20^. A study of 101 Chinese male IPF patients versus 53 controls matched for age and smoking history found that patients had significantly lower free testosterone (freeT; 210±8 pmol/l versus 281±13 pmol/l, p<0.001), and that leukocyte telomere length (LTL) was positively associated with freeT, independent of age and rare variants in telomerase genes (P=0.001)^21^.

Approximately 25-60% of idiopathic PF patients have LTL below the 10^th^ percentile^22-26^, suggesting telomere-mediated degenerative disease, and the proportion is much higher amongst familial cases with identified variants^24^. We hypothesised that sex hormones protect against onset and progression of PF in such cases. Using UK Biobank (UKBB) data for IPF cases and controls we therefore investigated associations between blood LTL ^27^, sex hormone concentrations and IPF frequency and progression, using observational regression, survival analyses and MR methods to explore causality.

## Methods

### Study design and participants

UK Biobank (UKBB) contains observational and genetic data from ∼500,000 adult volunteers aged 37-73 years, collected across the UK between 1/3/2006 and 31/3/2010^28^. Self-reported menopause age, hormone replacement therapy (HRT), clinical follow-up via hospital record linkage and LTL (available for 474,000 participants^27^) were analysed. A cohort of 379,708 unrelated participants of European descent was derived using ancestral principal components. This included 1133 IPF cases (defined by primary or secondary Hospital Episode Statistics (HES) ICD10 code of J84.1) and 378,575 controls. Details of the derivation with genotyping, and ethical approvals have been described previously^10^.

### Observational Associations

Key demographics were compared using logistic regression models in Stata 16.1, adjusting throughout for confounders of sex hormone concentrations (age, ever having smoked, BMI, social deprivation, and fasting time where hormone levels were used), unless stated otherwise. The majority of females in UKBB are postmenopausal with oestrogen levels below the limit of detection, therefore associations with premature ovarian failure or primary ovarian insufficiency (defined here as menopause <40years), early menopause (age<45years), hysterectomy, bilateral oophorectomy and prescription/administration of HRT (oestrogen only and oestrogen + progesterone) were investigated using our derived variables^29,30^. Associations between hormone change milestones (i.e., age of natural or surgical menopause) and age of PF diagnosis and all-cause death were also considered. HRT data were analysed by start date (pre- or post-2002) to avoid additional confounding following a 2002 publication on cancer risk which significantly impacted HRT prescribing practice in the UK^31^. Where specified, cases and controls were age-matched using ‘calipmatch’^32^. Total testosterone, SHBG and serum albumin were measured at recruitment; bioavailable and free testosterone levels were derived using SHBG/albumin concentrations^33,34^ and inverse normalised for use in regression analyses^35^.

Associations between IPF and its comorbidities were investigated with LTL as the exposure (using z-standardised natural logarithmic values of measured LTL^27^). Associations of age-adjusted LTL with the key confounders known to affect sex hormone concentrations were computed, and links (adjusted for confounders) were examined between hormone-related medical conditions with LTL as the outcome. Only data from post-menopausal women (over age 55 at registration) were used to avoid selection bias due to the UKBB age profile.

Cox proportional hazards regression was used to explore onset of, and survival with, IPF in males and females separately, using LTL, sex hormone measurements and covariates. Start time for onset was UKBB registration until date of diagnosis, or from date of diagnosis until HES-recorded death. Survival was censored to the HES data cut-off date (27/03/2017). Proportional hazards assumptions were tested based on Schoenfield residuals.

### Mendelian Randomisation (MR)

MR is a statistical approach that uses natural genetic variation and its influence on phenotype to provide information about an exposure and an outcome^36^. Since genetic variants are randomly assigned at conception, confounding and reverse causality are unlikely, making the genetic approach akin to a randomised control trial. Genetic variants robustly associated with sex hormone related outcomes (e.g. early menopause age <45 years) derived from genome wide association studies were used to create genetic instruments for early menopause in women^37^, total and free testosterone concentrations in men^38^ and SHBG concentrations in both^38^ (see Supplement for details). The genetic risk score (GRS) for SHBG (‘male SHBG cluster’)^38^ has been described previously as a genetic instrument with primary SHBG-increasing effects, and secondary divergent effects on total (higher) and bioavailable testosterone (lower), consistent with the known hormone-regulatory role of SHBG. For menopause age, two GRS were tested, the first containing 56 SNPs derived from meta-analyses independent of UKBB data^37^ and the second containing 290 SNPs based on meta-analyses including UKBB data but with effect estimates generated in GWAS excluding UKBB^39^, thus reducing the influence of Winner’s curse ^40^. GRS for telomere length was previously reported^10^. Association between IPF prevalence, age of diagnosis, age of death and the GRS in the unrelated data sets of 204,736 females and 174,972 males was investigated using one-sample MR with logistic regression models. Ancestral principal components (as previously described^41^) were included as covariates in the analysis to control for residual population structure; results were adjusted for UKBB assessment centre and array chip. Where numbers were very low for MR (i.e. when investigating links with age of diagnosis [N=402] and age of death [N=199] amongst female IPF cases), the cohorts were divided into deciles and MR performed using the lowest and highest deciles, with corresponding sample sizes, n=31 and n=29 respectively (reduced by missing data).

## Results

### Demographics

The well-established association between IPF incidence and male sex was replicated in this UKBB data set, after confounder adjustments: OR = 1.70 [95%CI: 1.50-1.93], P<1×10^−15^. The association of male sex with death amongst the 1,133 unrelated IPF cases was also consistent with previous reports ^19^: OR=1.58 [95%CI: 1.22-2.06], P=6.3×10^−4^. Established associations were noted between IPF and a range of demographic and environmental variables such as age and smoking, plus newly reported strong associations with the two variables relating to leukocyte telomere length (LTL): adjusted T/S ratio and Z-adjusted T/S log (see Supplement), (Tables 1 & 2). A 1-SD decrease in LTL increased the odds of IPF by 1.58 (95%CI: 1.49-1.67], P=1×10^−57^.

**Table 1:**
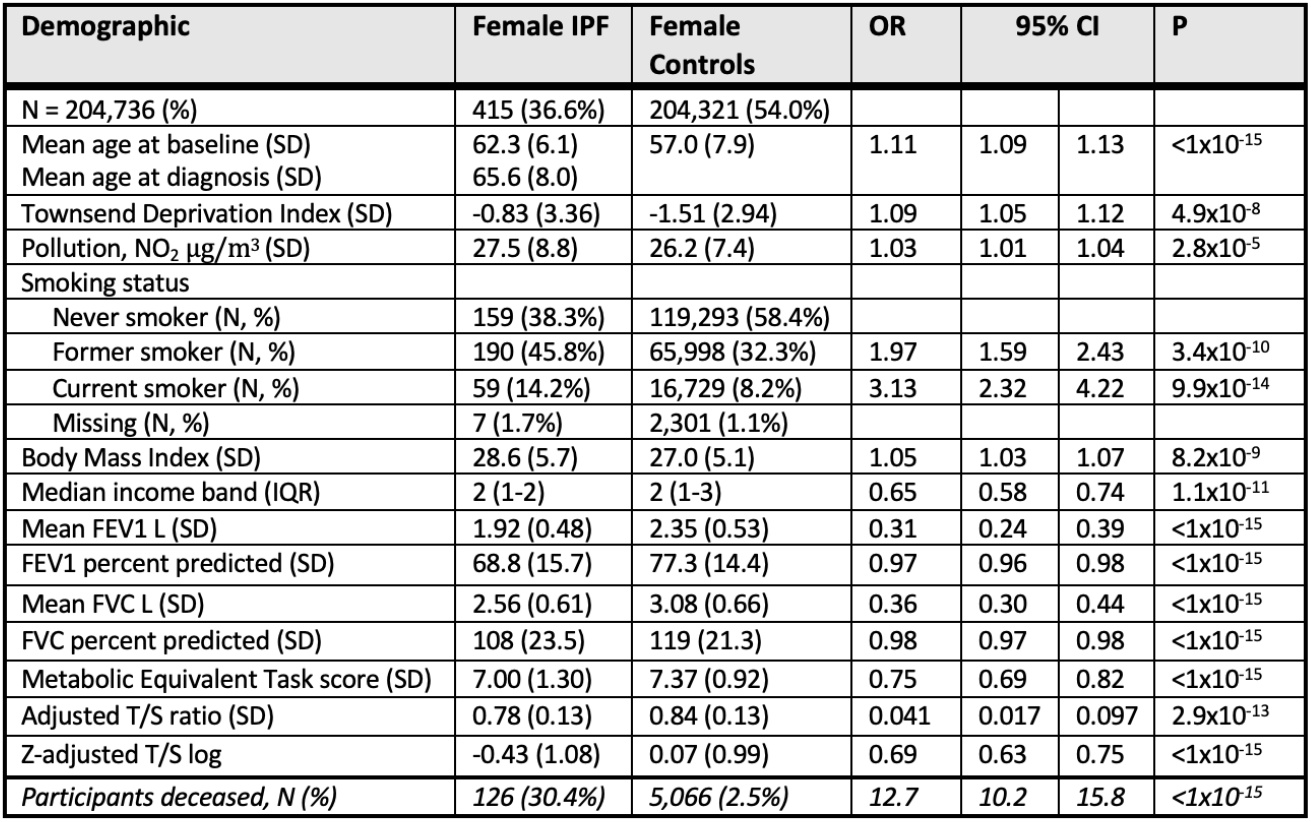
Demographics for unrelated female idiopathic pulmonary fibrosis cases and controls of European ancestry in UK Biobank derived using logistic regression analyses. Odds ratios show the extent to which the exposures predicted prevalence of IPF, except for ‘participants deceased’, which shows the risk ratio for death with IPF. (Odds ratios and p values are adjusted for age, other than percent predicted spirometry which already includes adjustment). Income bands indicate average household income: (1) <£18000, (2) £18000-£30999, (3) £31000-51999, (4) £52000-£100000 and (5)>£100000.

**Table 2:**
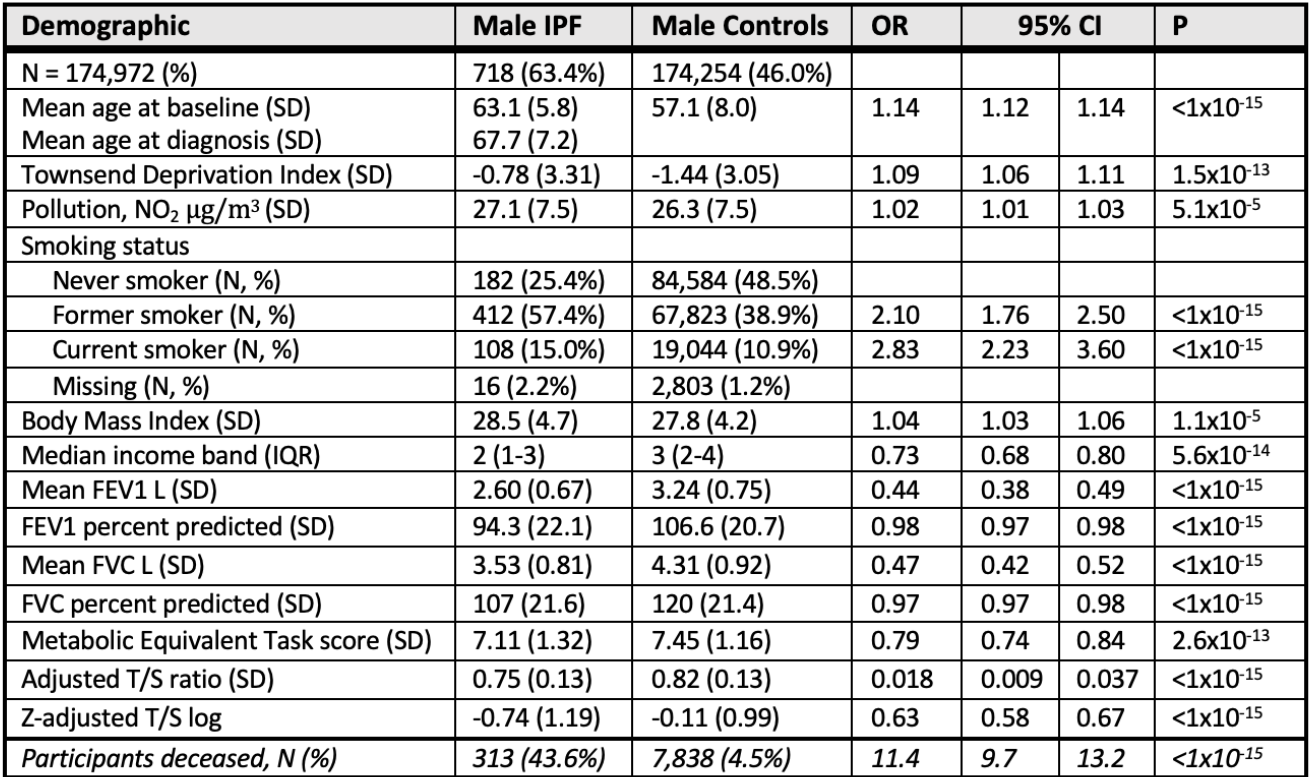
Demographics for unrelated male idiopathic pulmonary fibrosis cases and controls of European ancestry in UK Biobank derived using logistic regression analyses. Odds ratios show the extent to which the exposures predicted prevalence of IPF, except for ‘participants deceased’, which shows the risk ratio for death with IPF. (Odds ratios and p values are adjusted for age, other than percent predicted spirometry which already includes adjustment). Income bands indicate average household income: (1) <£18000, (2) £18000-£30999, (3) £31000-51999, (4) £52000-£100000 and (5)>£100000.

### Observational associations of IPF and its comorbidities with measured LTL

Building on previously reported findings for multiple other diseases in both sexes together^42,43^, IPF prevalence showed strong associations with LTL for both males (OR=0.63, P=2.2×10^−38^) and females (OR=0.69, P=8.9×10^−16^), with odds ratios much lower than those for other reported associated comorbidities of IPF such as coronary artery disease and type II diabetes (Figure 1, Supplement:S1). A similar, slightly weaker association was also found for a broader clinical definition of interstitial lung disease (ILD). Interestingly, the age-adjusted associations differed between sexes for some of these outcomes, including death.

**Figure 1:**
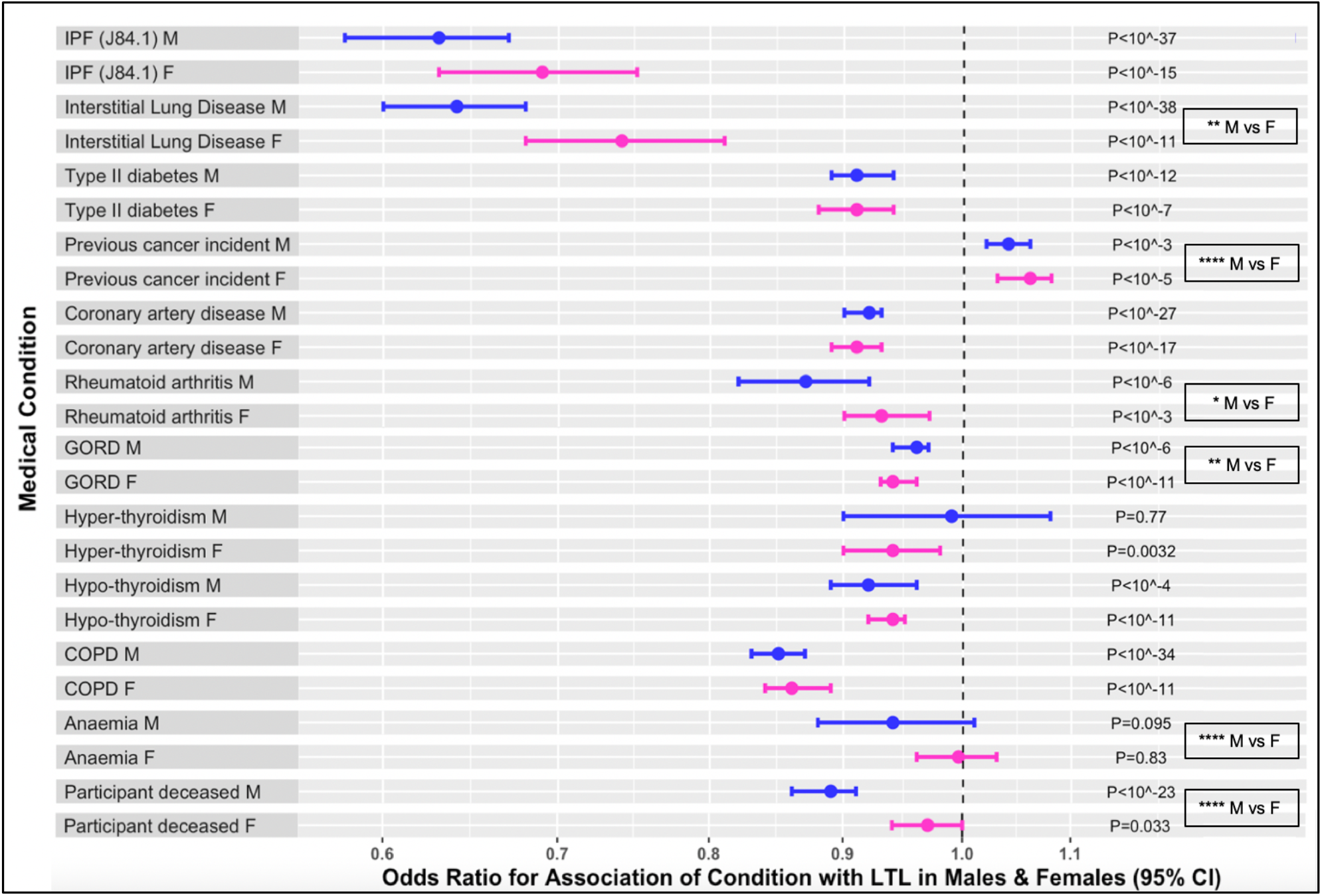
**Observational associations between prevalence of IPF and its related medical conditions with leukocyte telomere length** using z-adjusted loge T/S amongst female and male participants of white European ethnicity, with no adjustments for confounders other than age. Mean strengths of association are shown with 95% error bars. The hashed line indicates no change in odds of disease with changes in telomere length and the points furthest below or above this line show strongest decreased or increased odds of disease as LTL increases. The strongest disease associations with LTL were seen for IPF and more general interstitial lung disease. For some outcomes including death, the age-adjusted associations differed between sexes. P values indicate the significance of the association in each case. Stars highlight differences in strength of the associations for males and females as follows: *P ≤ 0.05, **P ≤ 0.01, *** P ≤ 0.001, *** P ≤ 0.0001. Telomere associations are shown throughout using pink for females and blue for males, for ease of intuitive assimilation, with hormone associations shown throughout using orange for females and turquoise for males.

### Association between factors affecting sex hormones and LTL

Associations were found between shorter LTL and the potential confounders we identified as affecting sex hormone levels (smoking, BMI, social deprivation and fasting duration) (Figure 2, Supplement: S2), with large effect sizes for current and male smokers. Importantly for this study, and as reported elsewhere^27^, the negative effect of age on LTL for females over the age of 55 (i.e. post-menopause) is greater than for younger women and similar to that of males whereas for younger women it is significantly better than for men.

**Figure 2:**
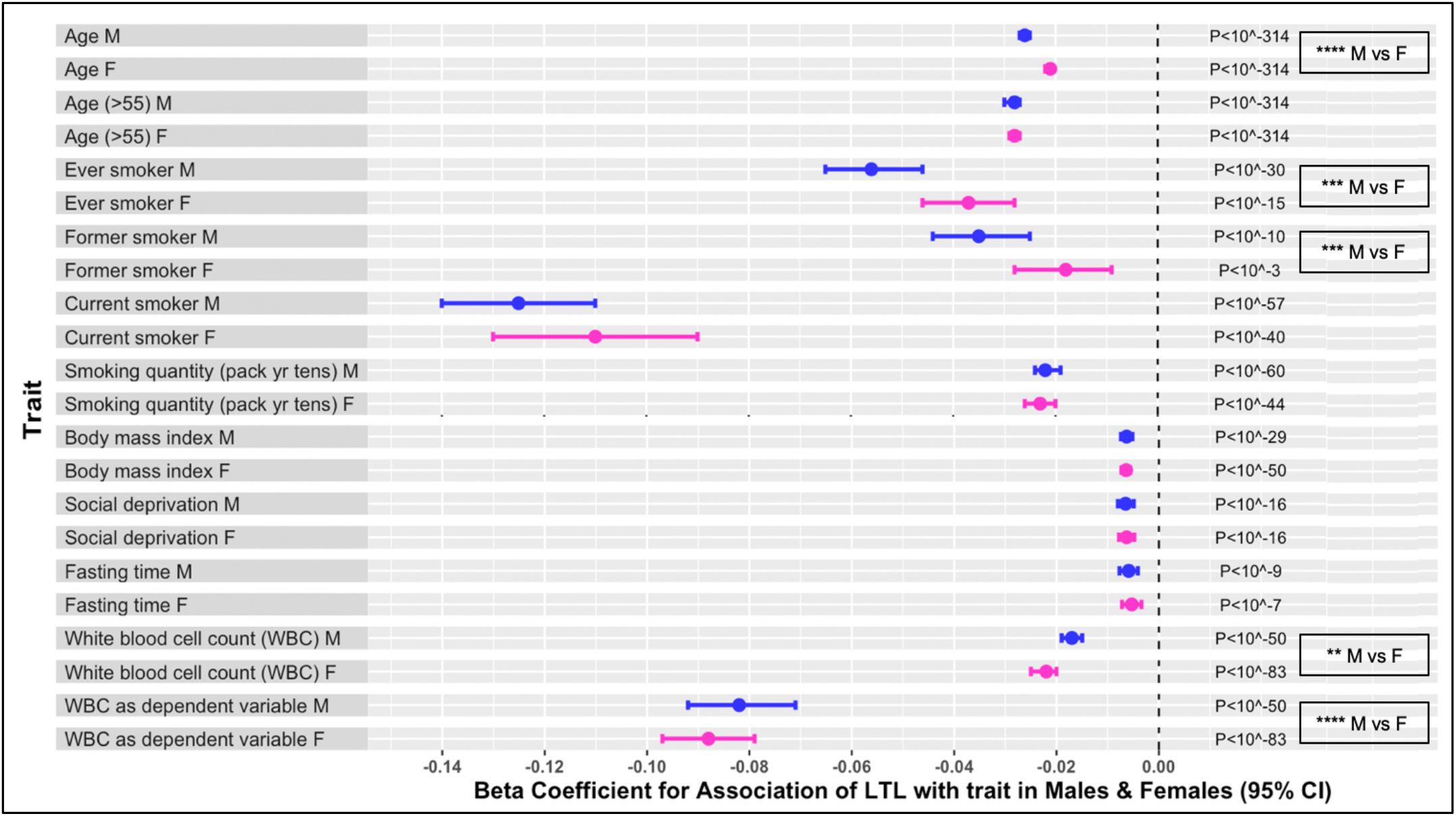
**Observational associations of leukocyte telomere length using z-adjusted loge T/S with key characteristics, in particular those known to be associated with sex hormone levels, amongst female and male participants** of white European ethnicity, with no adjustments for confounders other than age. Mean strengths of association are shown with 95% error bars. The hashed line indicates no change in mean telomere length with the trait and the points furthest below this line show strongest odds of decreased LTL as the trait increases. Large effect sizes are seen for current and male smokers and the impact of age is smaller for pre-menopausal women than for older women and for men. Stars highlight differences in the means between males and females as follows: *P ≤ 0.05, **P ≤ 0.01, *** P ≤ 0.001, *** P ≤ 0.0001. White blood cell count was included here for consideration of its role as a possible confounder or dependent variable in telomere biology disorders (although the LTL measurement is independent of cell count).

### Observational associations between PF and sex hormones

Amongst female IPF cases and age-matched controls (match ratio 1:200), IPF prevalence was associated with conditions of sex hormone depletion, after adjusting for the key confounders for menopause age^30^ (Figure 3A, Supplement: S3a). The strongest association was for premature ovarian failure (menopause <40yrs); OR=3.24 [95%CI:1.85-5.68], P=4.2×10^−5^. IPF prevalence was also negatively associated with age of natural menopause: OR=0.95 [95%CI:0.92-0.97], P=5.9×10^−5^. Mean age of natural menopause was lower amongst IPF cases than age-matched controls: 48.9 (± 5.6 SD) years for IPF cases (N=213) vs 50.2 (± 4.5 SD) years for controls, *β* coefficient = -1.33 [95%CI: -1.94, -0.72], P=2.0×10^−5^. The mean gap between age of natural/surgical menopause and age of IPF diagnosis was 18.7 (±8.8 SD) years, with only 5 cases being diagnosed prior to menopause. There was a significant association between natural or surgical menopause age and age of diagnosis for N=264 IPF cases (reduced by missing covariate data): *β* coefficient = 0.27 [95%CI:0.14-0.41], P=8.9×10^−5^ (Figure 3B). Similarly, there was an association between menopause age and age of death for N=80 cases, with on average a 10-year increase in menopause age corresponding to a 2.3 year longer life: *β* coefficient =0.22 [95%CI:0.024-0.41], P=0.028.

**Figure 3:**
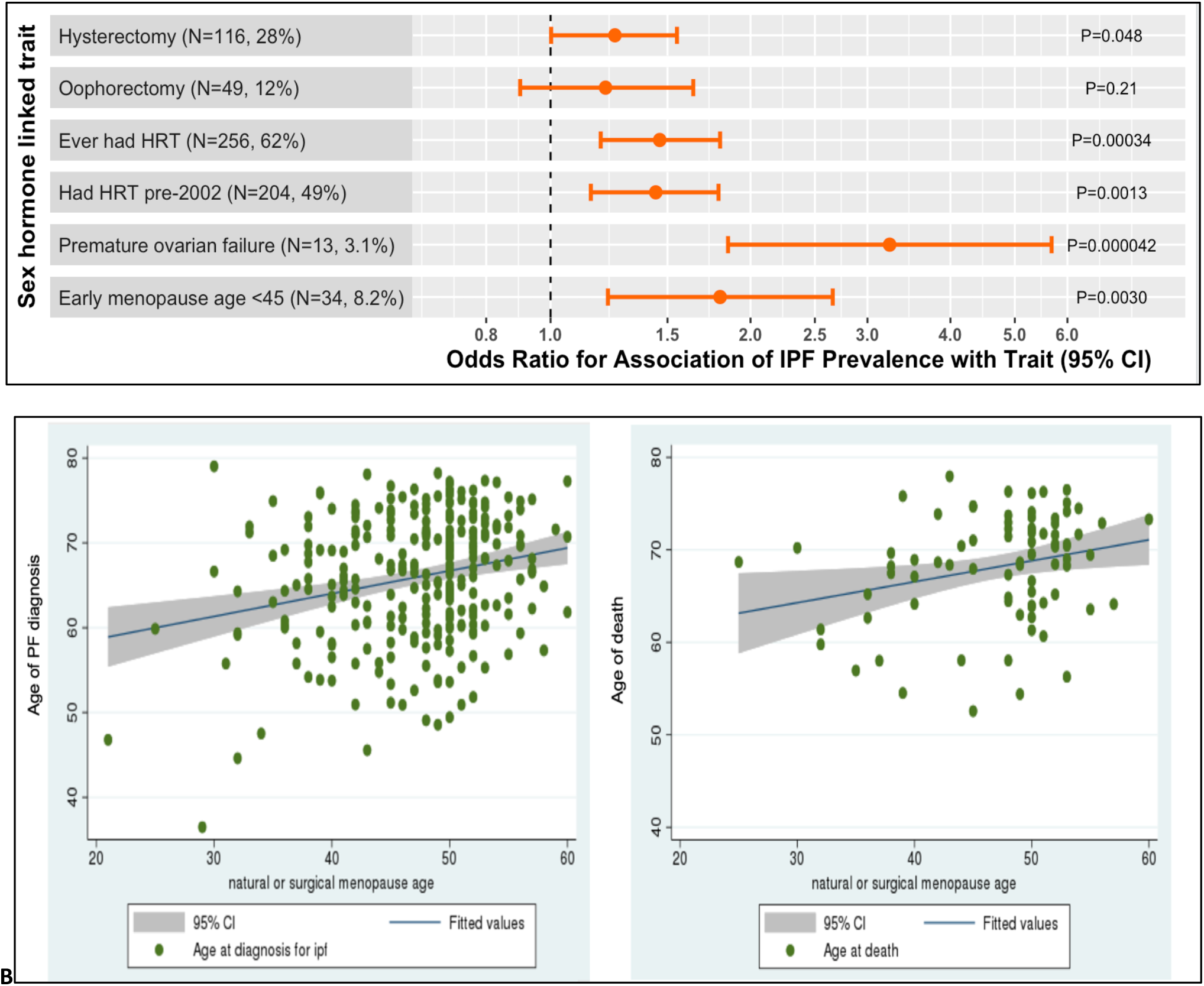
Associations of IPF prevalence and disease parameters with sex hormones amongst unrelated females of European ancestry in UK Biobank; (A) Associations of IPF prevalence with sex hormone related conditions for a female cohort of IPF cases and controls age-matched in the ratio 1:200 (to overcome confounding due to the change in prescribing policy for HRT in 2002). Odds ratios show the extent to which IPF is associated with each trait. Mean odds of association with 95% error bars are shown with the hashed line indicating no association. Analyses are adjusted for age, ever smoking, BMI and social deprivation. The number of cases included in each analysis varies due to missing data for the variables and covariates involved. (B) Association between IPF disease parameters (age of diagnosis and age of death respectively) and natural or surgical menopause age in female IPF patients (each point on the graph represents an individual IPF case with age of diagnosis or death data).

For male participants who later developed IPF, mean fractions of bioavailable testosterone and free testosterone were within normal range at registration but were lower than for age-matched controls (match ratio 1:100): bioavailable testosterone for cases was 39.6% (± 9.6 SD) compared with 42.3% (± 9.7 SD) for controls (β=-3.04, P=3×10^−16^) and free testosterone was 1.66% (± 0.40 SD) compared with 1.73% (± 0.39 SD) for controls (β= -0.081, P=5×10^−8^). Mean haemoglobin concentration for existing male IPF cases at registration was within normal range, at 14.5g/dL (± 1.6 SD).

SHBG controls sex hormone bioavailability, with higher concentrations reducing the bioavailable and free testosterone *fractions* and raising the total testosterone concentration (Figure 4A-C, Supplement S5). After covariate adjustments, IPF prevalence in males was associated positively with SHBG, OR=1.35 [95%CI: 1.23-1.47], P=2.6×10^−11^ per 1SD higher concentration at registration. IPF was associated negatively with bioavailable testosterone percentage: OR=0.69 [95%CI: 0.63-0.76], P=5.6×10^−16^ per 1SD increase (Supplement: S5), even though the majority of cases (612/718 males) were incident, with mean time to diagnosis = 4.99 years ± 2.32 SD. A weak association was also found amongst females: OR=0.85 [95%CI: 0.73-0.97], P=0.02. further observational associations between IPF and hormone level parameters are reported in the supplement.

**Figure 4:**
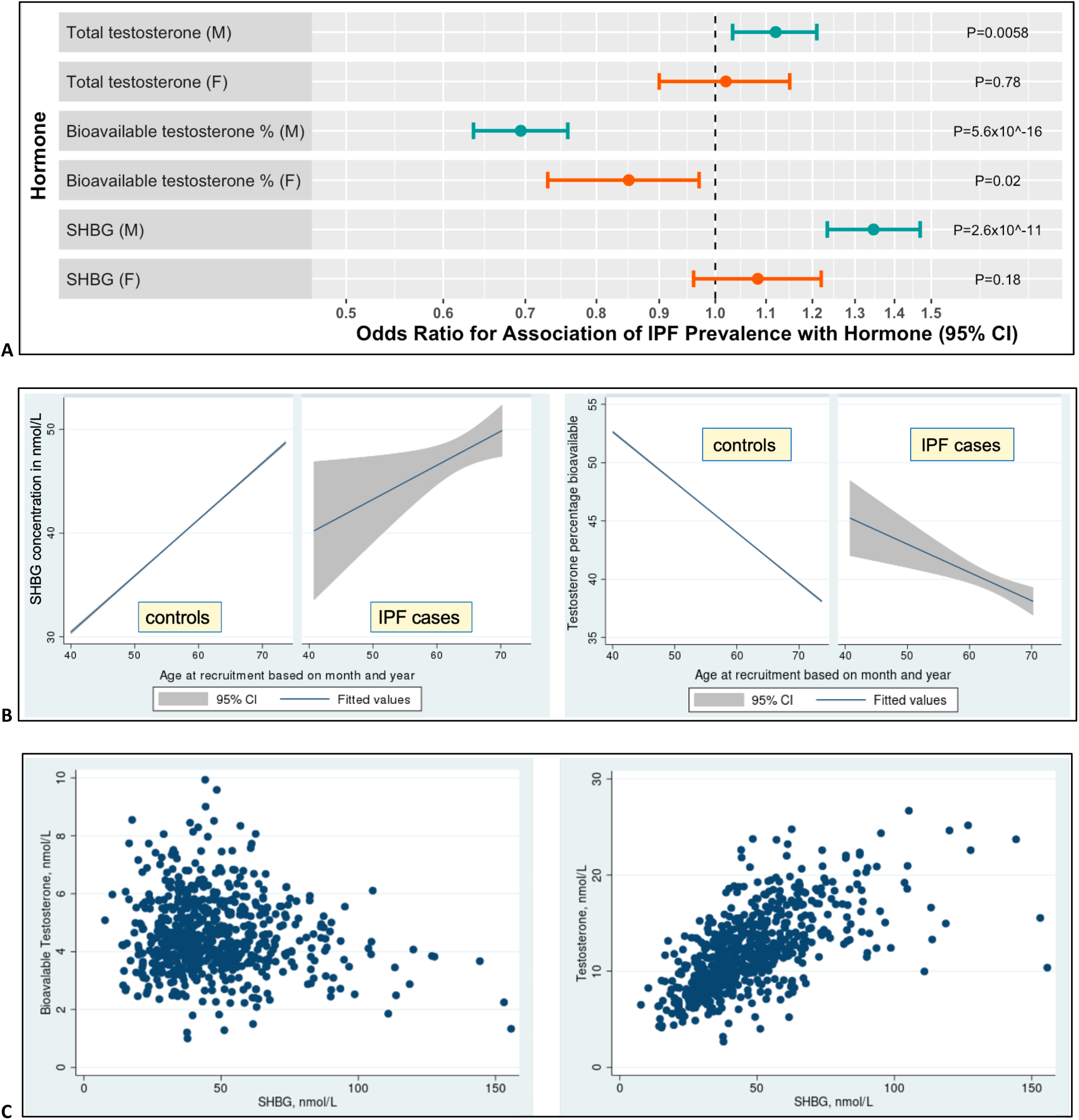
Associations of IPF prevalence with sex hormones amongst unrelated females and males of European ancestry in UK Biobank; (A) Odds of IPF prevalence amongst male and female IPF cases and controls for a 1SD (standard deviation) increase in sex hormone levels. Analyses are adjusted for age, ever smoking, BMI, social deprivation and fasting time. (B) Mean and 95% CI concentrations of SHBG and of bioavailable testosterone with age at recruitment for male controls and IPF cases (for controls, the 95% lines are very similar to the mean due to high numbers so not visible in the plot). (C) Scatter plots showing how bioavailable and total testosterone concentrations vary with SHBG in IPF (each point on the graph represents an individual IPF case). Sex hormone associations are shown throughout using orange for females and turquoise for males.

In summary, the data shows consistent multi-faceted evidence of PF prevalence linked with reduced sex hormone concentrations.

### LTL and sex hormones

The above conditions relating to sex hormone levels were also associated with shorter telomeres (Figure 5A, Supplement: S6A), with a striking association with early menopause: *β* = -0.087 [95%CI:0.112-0.61], P=2.9×10^−11^. There was no observed association of hysterectomy and oophorectomy with *shorter* telomeres, likely because the indications for surgery are diverse but typically included as part of cancer treatment. LTL was also positively associated with natural menopause age: *β* =0.0080 [95%CI:0.0065-0.0094], P=6.6×10^−27^.

**Figure 5:**
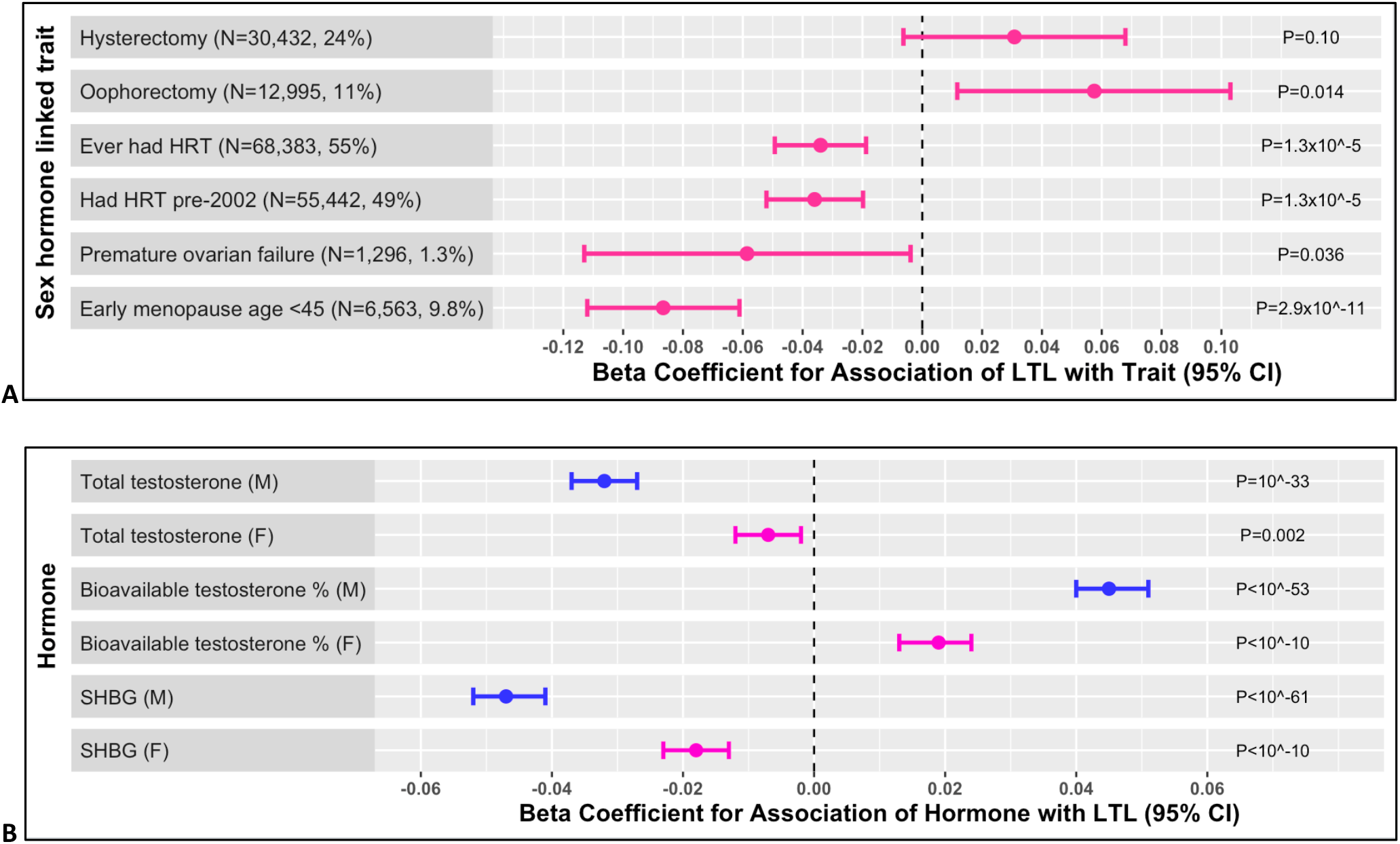
Associations of leukocyte telomere length with sex hormones amongst unrelated individuals of European ancestry in UK Biobank. Mean strengths of association are shown with 95% error bars.The hashed line indicates no change in association of telomere length with sex-hormone linked trait or concentration and the points furthest below or above this line show strongest odds of shorter or longer LTL associated with the trait or hormone concentration. **(A)** Associations of Z-adjusted LTL Log with sex hormone related conditions in females over the age of 55 at registration, showing that both having been prescribed HRT and having had early menopause (age <45 years) are associated with shorter telomeres. Beta coefficients show how LTL as the outcome variable is associated with each trait. Analyses are adjusted for age, ever smoking, BMI and social deprivation. The number of cases included in each analysis varies due to missing data for the variables involved. **(B)** Associations of Z-adjusted LTL Log with sex hormones measured at registration in males and females, showing strong negative associations of LTL with total testosterone and SHBG and positive associations with bioavailable testosterone in males. Analyses were adjusted for age, ever smoking, BMI, social deprivation and fasting time.

Mirroring the associations with IPF, LTL was associated negatively with total testosterone and SHBG but positively with bioavailable testosterone, again with considerably stronger associations in males then females (Figure 5B, Supplement: S6b).

### Sex hormones and disease progression

Multiple cox proportional hazard regression analyses for PF onset and survival showed lower hazard ratios linked with increasing LTL and sex hormone levels. Slower disease progression was linked with both later menopause and having received HRT for females and with higher bioavailable testosterone concentrations for males (see supplemental Results). This suggests that for patients with PF, restored hormone levels could be important for survival.

### Genetic assessment of causality

Evidence of a causal link between our SHBG GRS, ‘male SHBG cluster’ and telomere length was found amongst all UKBB participants: *β* =-0.00085 [95%CI:-0.0012,-0.0052], P=6.5×10^−7^ (N=365,075; adjusting for principal components, test centre, chip, fasting time and age), suggesting that higher SHBG leads to shorter telomeres and could therefore be implicated in PF causality. This GRS has been described previously and represents a genetic instrument with primary SHBG-increasing effects, and secondary divergent effects on total (higher) and bioavailable testosterone (lower) that are consistent with the known hormone-regulatory role of SHBG.

Using our telomere length GRS^10^ (which is strongly associated with measured LTL: R^2^ =0.009, F = 3411), there also was evidence suggesting that SHBG levels (single inverse normalised) are affected by shorter telomeres in men: *β* =0.011 [95%CI:0.008-0.014], P=7.9×10^−12^. Amongst women, there is weaker evidence of a link: *β* =0.0038 [95%CI:0.009-0.0067], P=0.010. Thus, in men, short telomeres appear to cause higher SHBG levels which would reduce the bioavailable testosterone further. This finding supports the possibility of a detrimental feedback loop for high SHBG and short telomeres amongst males with IPF. If a more stringent p value of p<0.001 is used to allow for multiple testing with 6 different hormone genetic risk scores, the above results in males remain robust. Additional MR analyses hinted at causality behind our reported observations but were limited by IPF case numbers that are too low for this type of analysis (see Supplement).

Thus, our genetic analysis provides evidence suggesting a bi-directional causal role in IPF for increased SHBG levels and short telomeres in males and some indications of a causal role for short telomeres and sex hormone insufficiency in females. Future large-scale genetic studies will enable further analyses of these links.

## Discussion

Observational analyses in UKBB data have shown evidence of multiple consistent associations between pulmonary fibrosis and lower sex hormone levels in males and females. These findings were underpinned by links between PF and short LTL which were then mirrored in relationships between LTL and hormone levels. Genetic causal analyses provided evidence of a causal interaction between SHBG and telomere length, most important in males where it appears to be bi-directional. Taken together, these findings support our hypothesis that high-normal sex hormone concentrations could be protective in PF, whereas low-normal sex hormone concentrations could be detrimental.

Recent findings have reported a strong observational association of longer telomeres with menopause age and both observational and MR associations between longer telomeres and lower SHBG and testosterone (males)^42^. Our results build on these, by highlighting the strength of association with LTL of IPF and ILD and through carrying out separate analyses in males and females to elicit important gender differences.

Reports suggest that sex hormones play a regulatory role in expression of TERT, with oestrogen receptors appearing to act directly on the *TERT* gene promoter to upregulate telomerase^44,45,46^ and androgens acting indirectly via a range of mechanisms including (but not limited to) aromatisation to oestrogen ^47-49,50,51^. Of particular interest, in cell culture of peripheral blood lymphocytes from patients heterozygous for telomerase mutations, low baseline telomerase levels were restored to normal by androgen exposure^49^. Amongst healthy women, long-term hormone replacement therapy (HRT) correlates with longer telomeres^52^.

Evidence for the potential therapeutic role of sex hormones in PF exists in the treatment of 27 TBD patients (15 female, median age 41 years and age range 17-66), with the synthetic androgen danazol ^53^. Whilst caution in interpretation is required as danazol is a weak androgen, weak progestogen, weak antigonadotrophin, weak steroidogenesis inhibitor and a functional antioestrogen, recruitment was halted early because 11/12 patients had an unanticipated gain in telomere length at 24 months. PF scores based on CT ^54^ and lung function DLCO measurements were stable during the 2 years of treatment for all patients but one. Due to side effects, several trials at lower doses are now underway^55-57^. In a dramatic recent case study, a 42 year old female PF patient due for lung transplant recovered after 18 months on danazol ^58^.

However, adopting a therapeutic approach to supplement sex hormone concentrations above physiological requirements requires key safety considerations and monitoring implications. Supraphysiological testosterone administration stimulates erythrocytosis and so causes secondary polycythaemia and hyperviscocity, which can increase the risk of cardio- and cerebrovascular events. Women beyond the age of natural menopause are only recommended to take HRT for 5 years to minimise the risk of stimulated breast cancer and increased thromboembolic events. However, as sex steroid concentrations were lower than age-matched controls, supplementing to higher concentrations within the normal range but not above could yield disease benefit in IPF but avoid at least some of the side effects. Similarly, upregulation of telomerase carries risks. Although overt TERT expression is insufficient to cause malignant transformation, its presence can promote unchecked proliferation; over-expression of telomerase is detected in around 90% of human cancers ^15^. Shorter telomeres are generally associated with mortality^42,43^, yet we found this to be the case for men only (Figure 2), probably because a higher proportion of female deaths are caused by cancer, which is associated with longer telomeres (in UKBB, 68% of deaths amongst the top 20 causes for females were cancer-related whereas for males, this figure was 47%^59^).

Our study is limited by the lack of extensive clinical characterisation of cases (although these have shown good consistency with replication cohorts previously^10^), and by the healthy volunteer effect amongst research participants. The MR studies amongst IPF cases are currently underpowered and replication in large clinical cohorts would be valuable. Some of the key variables (e.g. menopause age) are self-reported and so may be impacted by recall bias.

In summary, these findings provide evidence that lower sex hormone concentrations are related to PF and we speculate that treatment to supplement sex hormone concentrations higher within the normal reference range but not above it, may delay or prevent onset of PF amongst at risk individuals (such as relatives of patients with familial PF caused by germline telomere defects) and slow disease progression amongst patients with short telomeres. If correct, this may offer a relatively low-cost (particularly when compared to novel agents for IPF), available, and well-tolerated treatment which could be explored in a future, randomised clinical trial powered for safety and disease outcomes in men and women.

## Data Availability

All data produced in the present work are contained in the manuscript

## Author Contributions

AD conceived the project, carried out the analyses, interpreted the results and drafted the manuscript; AM, KSR, RNB, ARW and SM contributed to the statistical analyses, MAG, JKP and AMR provided clinical guidance and interpretation; HA and JC provided patient representation; KL, and MAL provided supervisory support to AD; JT and CJS were the principal investigators for the study, with oversight of study design, analysis and interpretation, plus administration of access to UK Biobank. All authors contributed to drafting and revision of the manuscript. All authors read and approved of the final manuscript.

## Acknowledgements

This work was supported in part by grant MR/N0137941/1 for the GW4 BIOMED MRC DTP, awarded to the Universities of Bath, Bristol, Cardiff and Exeter from the Medical Research Council (MRC)/UKRI. KSR is supported by Cancer Research UK [grant number C18281/A29019]. JKP is supported by the UKRI Expanding Excellence in England award. JT is supported by an Academy of Medical Sciences (AMS) Springboard award, which is supported by the AMS, the Wellcome Trust, GCRF, the Government Department of Business, Energy and Industrial strategy, the British Heart Foundation and Diabetes UK [SBF004\1079]. CJS is supported by MRC project grants (MR/V002538/1 and MR/S002626/1). This research has been conducted using the UK Biobank Resource (applications 9072 and 44046).

