## Supplementary Material for "Study of the associations between short telomeres, sex hormones and pulmonary fibrosis"

### Methods: additional information

Details and derivation of the two leukocyte telomere length variables used, "Adjusted T/S ratio" and "Z-adjusted T/S log", (UK Biobank data fields 22191 and 22192 respectively) have been reported elsewhere[1] and are summarised here. The relative leucocyte telomere length ("T/S ratio", UK Biobank data field 22190) was measured using a validated quantitative PCR (polymerase chain reaction) assay and expressed as the ratio of the number of telomere repeats relative to a reference single-copy haemoglobin subunit beta gene. This ratio is proportional to an individual's average telomere length[2]

> "Adjusted T/S ratio": T/S ratio adjusted for operational and technical parameters (PCR machine, staff member, enzyme batch, primer batch, temperature, humidity, primer batch × PCR machine, primer batch × staff member, A260/A280 ratio of the DNA sample, and A260/A280 ratio squared)
>
> "Z-adjusted T/S log": adjusted T/S ratio which has been both $\log_e$-transformed to obtain a normal distribution and then Z-standardised using the distribution of all individuals with a telomere length measurement.

Details of the testosterone based genetic instruments[3] used for Mendelian randomisation causal analysis are as follows (where N is the number of SNPs):

> "Total testosterone-men" (N=231): Individually genome-wide significant SNPs for total testosterone in men, weighted by individual SNP beta estimate for total testosterone
>
> "Bioavailable testosterone – men" (N=125): Individually genome-wide significant SNPs for bioavailable testosterone in men, weighted by individual SNP beta estimate
>
> "Bioavailable testosterone – all" N=147: Individually genome-wide significant SNPs for bioavailable testosterone in men and women combined, weighted by individual SNP beta estimate
>
> "Male SHBG cluster" (N=362): formed from SNPs with primary effects on SHBG in men, and secondary effects on total and bioavailable testosterone. Each SNP in this genetic instrument is weighted by its effect from the BMI unadjusted SHBG analysis.

### Results: additional information

#### Observational associations between PF and sex hormones

Serum albumin concentration also promotes the bioavailability of testosterone; mean albumin concentrations at recruitment were lower for IPF cases than controls and IPF prevalence was associated with albumin level in both sexes combined: for a 1SD increase in albumin concentration, OR=0.65 [95%CI: 0.61-0.69], $p<1x10^{-38}$. Similar associations were evident in females and males. However, LTL was not linked with albumin levels in either sex.

#### Sex hormones and disease progression

Cox proportional hazards regression for time from registration to diagnosis of IPF amongst females showed hazard ratios of HR=0.69 [95%CI:0.63-0.75], $P=6.4x10^{-18}$ for LTL adjusted initially for age only. When added to this model, age of natural menopause had HR=0.97 [95%CI:0.94-0.996], P=0.028, suggesting that both longer telomeres and later menopause (or longer duration of higher sex hormone concentrations) are protective against developing IPF (Figure S1). These relationships persisted at P<0.05 in a model containing covariates (smoking, BMI and deprivation) (Table S7a).

Similar analyses for time from registration to death (any cause) with IPF amongst females showed HR=0.64 [95%CI:0.58-0.71], P=2.0x10$^{-17}$ for LTL (adjusted for age only) and HR=0.94 [95%CI:0.90-0.98], P=0.0076 for age of natural menopause (adjusted for age and LTL), suggesting that longer telomeres and later menopause are linked with survival in IPF (Figure S1B). The observed association with time to death was stronger than for time to diagnosis despite considerably smaller numbers (and again persisted at P<0.05 when confounders such as smoking status were included) (Table S7b).

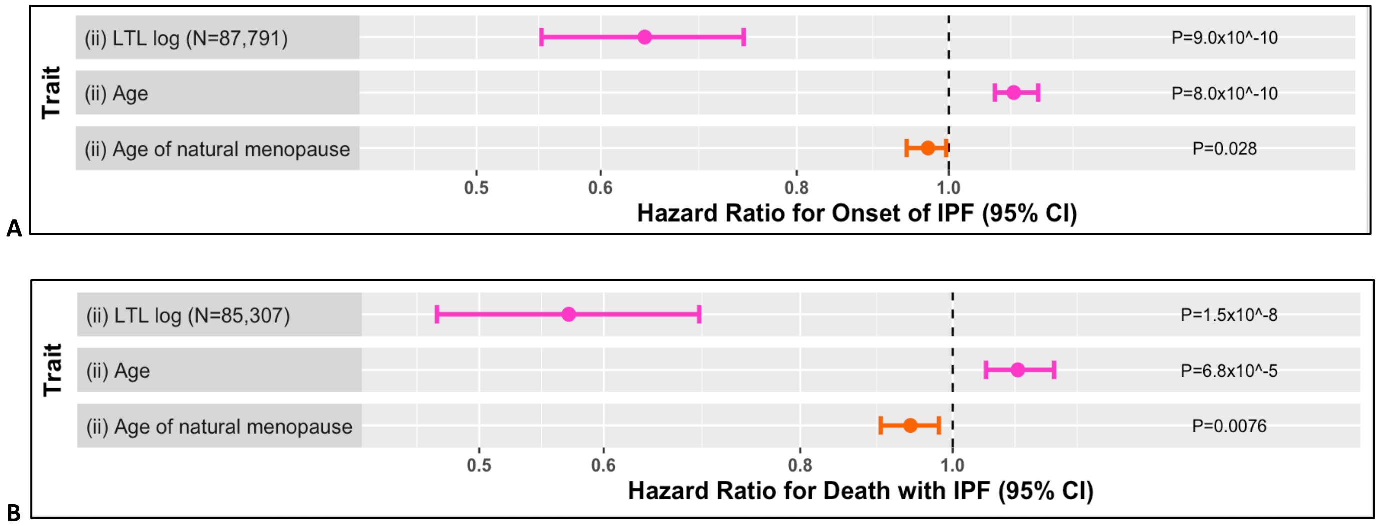


**Figure S1. Cox proportional regression results based on time from registration to IPF diagnosis and to IPF death with respect to LTL and age of menopause for females in UK Biobank. This plot shows the mean increase or decrease in hazard ratio (with 95% confidence interval) for each trait, with no impact on hazard shown by the hashed line. (A)** hazard ratio for LTL in a model for time from registration to IPF diagnosis, including age and age of natural menopause, **(B)** hazard ratio for LTL in a model for time from registration to death with IPF, including age and age of natural menopause. Both analyses were restricted to those who had natural menopause.

### Link between hormone replacement treatment and disease

Participants who later developed IPF were more likely to have received HRT than age-matched controls: OR=1.46 [95%CI:1.19-1.80], P=3.4x10$^{-4}$ (Table S3) and this result was attenuated to the null after controlling for natural menopause age, which is expected if HRT was prescribed to counter hormone insufficiency causing early menopause, as per normal clinical practice. Amongst cases and age-matched controls, the mean age of starting HRT was earlier amongst IPF cases: 45.7 vs 48.1years, and IPF prevalence was associated with younger age of requiring HRT: OR=0.93 [95%CI:0.91-0.96], P=9x10$^{-9}$. These findings suggest that increasing hormone insufficiency correlates with higher risk of developing IPF.

For those IPF cases who had received HRT, the age of IPF diagnosis was associated with both the treatment start age and end age (which were strongly correlated), but not with duration of treatment (Table S4, Figure S2), such that on average each additional 10-year increase in HRT start-age corresponded to a 4.5-year delay in disease onset. Similar weaker associations were seen with age of death. These findings suggest that the stages of disease are linked with declining hormone levels. LTL also correlated with age of starting HRT: $\beta$ = 0.07 [95%CI: 0.03, 0.10], p=0.001. There was 6.2% overlap between having received HRT and having had early menopause (which includes premature ovarian failure here).

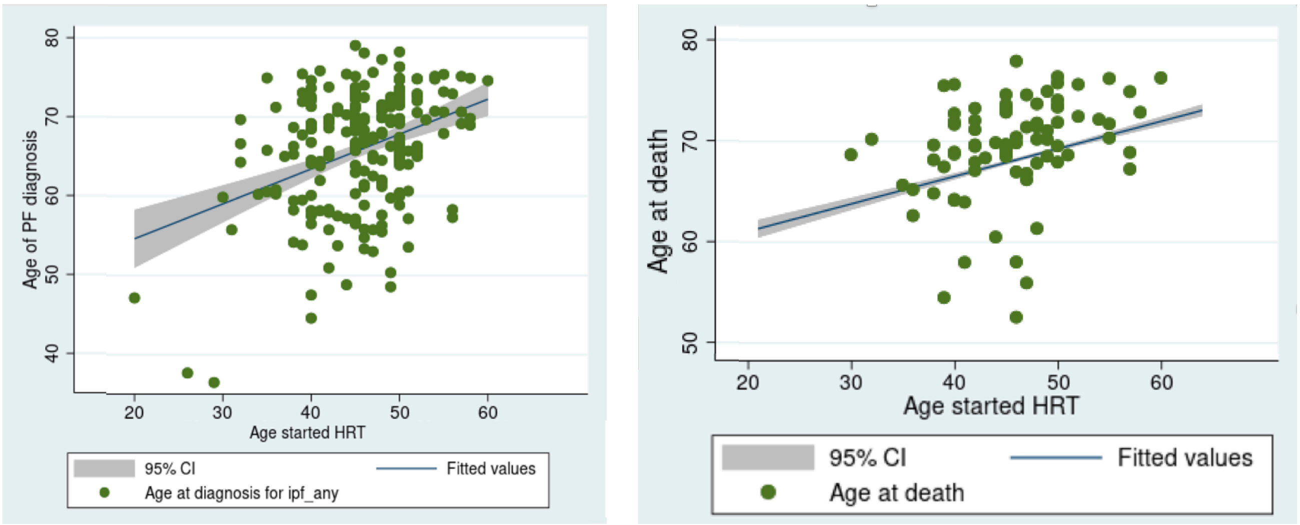


**A** **B**

**Figure S2. Association between IPF disease parameters and age started HRT for female IPF patients; (A)** age of diagnosis vs age of starting HRT, **(B)** age of death vs age or starting HRT (each point on the graph represents an individual IPF case with age of diagnosis or death data).

Cox proportional hazard analysis for time from IPF diagnosis to death amongst all post-menopausal cases (N=241) showed that HR=0.75 [95%CI:0.59-0.95], P=0.016 for LTL and HR=0.60 [95%CI:0.37-0.98], P=0.041 for having received HRT (in a model including sex hormone-related covariates), (Figure S3, Table S8), suggesting that both longer LTL and having received HRT were linked with slower decline. When menopause age was included as a covariate, the results for HRT were HR=0.62 [95%CI:0.37-1.04], P=0.071.

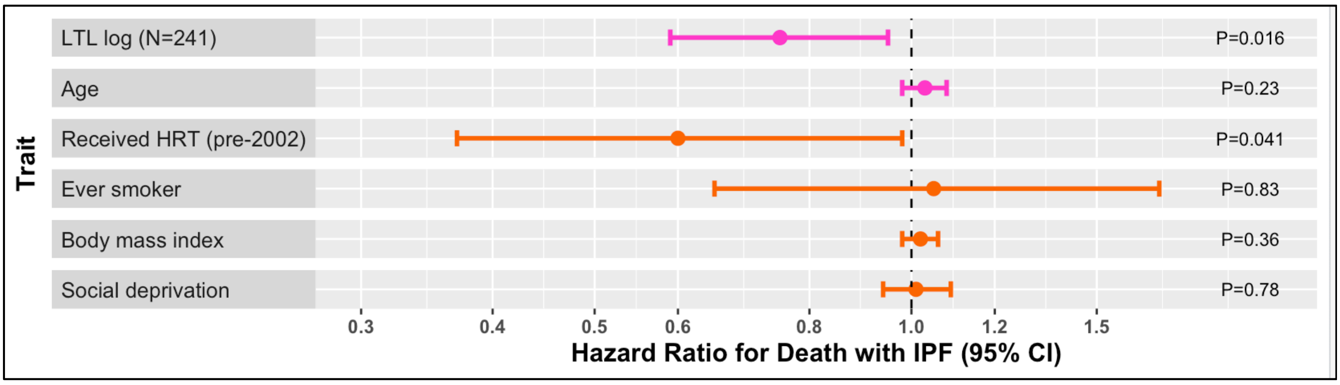


**Figure S3. Cox proportional regression analysis of time from diagnosis to death amongst female IPF cases in UK Biobank** (analysis was restricted to those participants who were post-menopausal to avoid age selection bias). This plot shows the mean increase or decrease in hazard ratio (with 95% confidence interval) for each trait, with no impact on hazard shown by the hashed line.

Amongst males, Cox proportional hazards regression for time from registration to diagnosis of IPF showed hazard ratios HR=0.61 [95%CI:0.57-0.66], P=$2.4 \times 10^{-39}$ for LTL adjusted for age only and HR=0.88 [95%CI:0.82-0.94], P=$3.6 \times 10^{-4}$ for bioavailable testosterone concentration in a model adjusted for both age and LTL, suggesting that both longer telomeres and higher bioavailable testosterone levels are protective against developing IPF (Figure S4A, Table S9b). In a similar model, SHBG concentration was positively associated with risk of diagnosis: HR=1.012[95%CI:1.007-1.016], P=$1.2 \times 10^{-7}$. When bioavailable and free testosterone levels were expressed as percentages of total testosterone, both were associated with risk of IPF diagnosis: HR=0.97 [95%CI:0.96-0.98], P=$8.6 \times 10^{-10}$ and HR=0.62 [95%CI:0.49-0.79], P=$1.1 \times 10^{-4}$ respectively.

These relationships persisted in models containing sex confounding covariates of sex hormone concentrations (Table S9a=i).

A
B

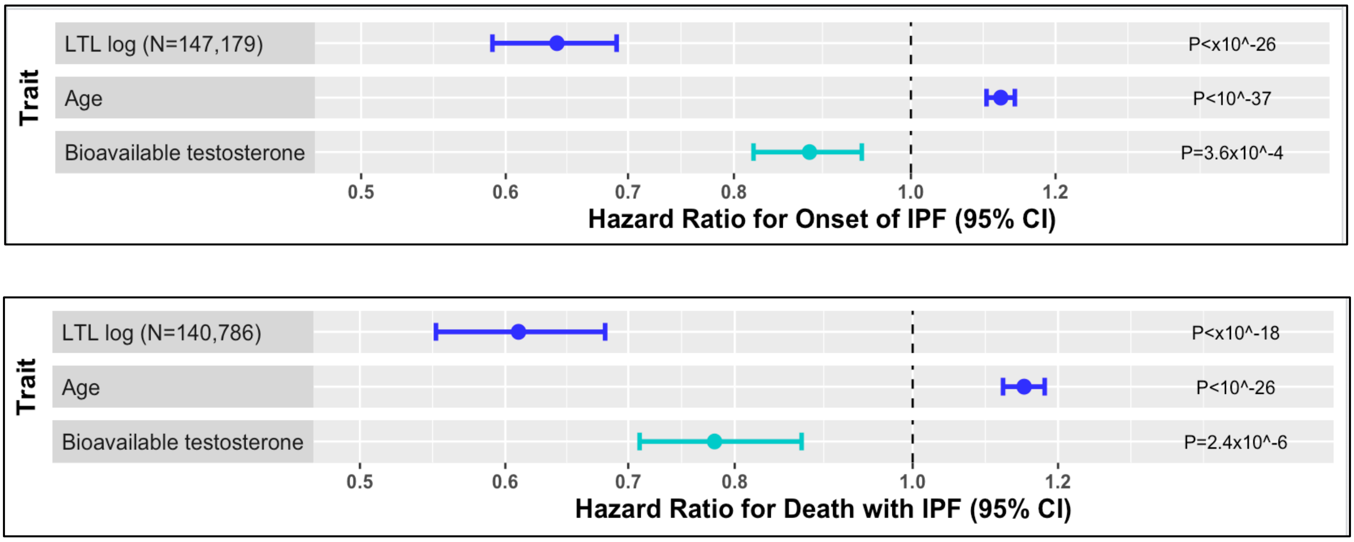


**Figure S4. Cox proportional regression results based on time from registration to IPF diagnosis and to IPF death with respect to LTL and bioavailable testosterone concentration at registration for males in UK Biobank. This plot shows the mean increase or decrease in hazard ratio (with 95% confidence interval) for each trait, with no impact on hazard shown by the hashed line.** **(S4A)** hazard ratio for LTL in a model for time from registration to IPF diagnosis, including age and bioavailable testosterone concentration, **(S4B)** hazard ratio for LTL in a model for time from registration to death with IPF, including age and bioavailable testosterone concentration.

Similar analyses for time from registration to death with IPF amongst males showed hazard ratios of HR=0.59 [95%CI:0.54-0.65], P=5.1x$10^{-28}$ for LTL (adjusted for age only) and HR=0.78 [95%CI:0.71-0.87], P=2.4x$10^{-6}$ for bioavailable testosterone concentration (adjusted for age and LTL), suggesting that longer telomeres and bioavailable testosterone levels are linked with survival in IPF (Figure S4B, Table S9e). When bioavailable and free testosterone levels were expressed as percentages of total testosterone, both were negatively associated with death from IPF: HR=0.96 [95%CI:0.95-0.97], P=1.4x$10^{-8}$ and HR=0.57 [95%CI:0.41-0.79], P=9.1x$10^{-4}$ respectively, suggesting that clinically measured lower testosterone levels are associated with IPF mortality. Higher SHBG levels were also associated with IPF mortality (Tables S9d-f). Again, the observed association with time to death was stronger than for time to diagnosis despite considerably smaller numbers.

Amongst male IPF cases with data for all covariates (N=578), Cox proportional hazards analysis of time from registration to death showed weak evidence of an association of bioavailable testosterone concentration with survival HR=0.89 [95%CI:0.81-0.98], P=0.021 (in a model including sex hormone related covariates, Figure S5). Similar models for free testosterone and SHBG did not reach significance (Tables S9g-i). This suggests that for patients with IPF, bioavailable testosterone concentration could be important for survival.

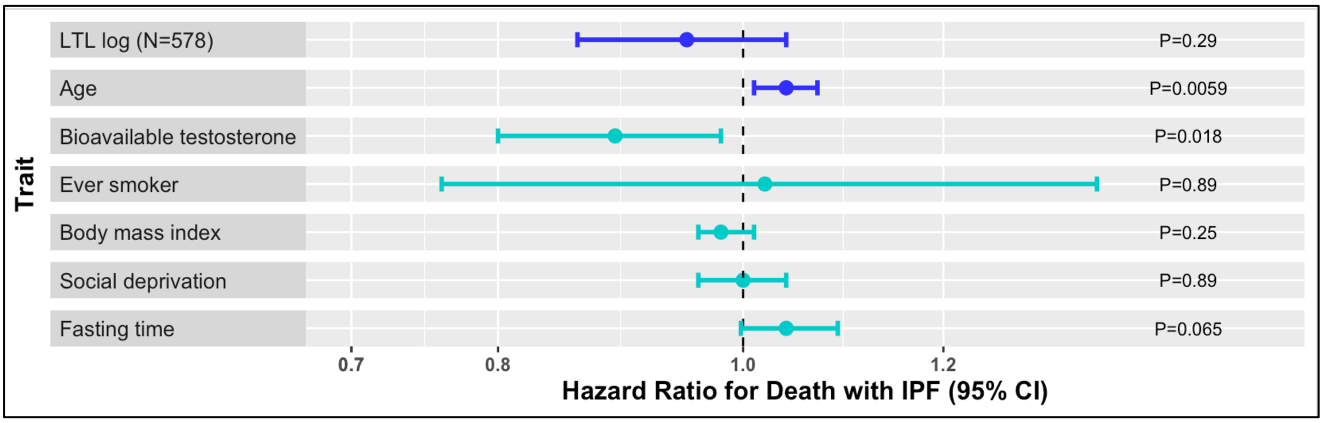


**Figure S5.** Cox proportional regression analysis of time from registration (at which biomarkers were measured) to death amongst male IPF cases in UK Biobank. **This plot shows the mean increase or decrease in hazard ratio (with 95% confidence interval) for each trait, with no impact on hazard shown by the hashed line.**

### Genetic assessment of causality: additional information

Despite IPF case numbers being too low for meaningful MR analyses, results in this cohort suggested at causality behind our reported observations. In our female IPF cases dataset, we looked for evidence of a causal link between a version of our 290 SNP genetic risk score for early menopause based on effects in non-UKBB strata and age of death for IPF cases in this cohort who died before September 2020 using the lowest and highest deciles. Sample size was very small (n=29) and no evidence was found (OR=1.19 [95%CI: 0.96, 1.47], P=0.12. However, as a sensitivity study using a 56 SNP GRS for early menopause that is independent of UKBB [4] and age of death for IPF cases, a weak causal link was apparent: OR=1.80 [95%CI: 1.13-2.85], P=0.013 (n=29)]. In our male IPF dataset there was insufficient evidence of an association between our male bioavailable testosterone GRS and age of death $\beta$=0.10 [95%CI:-0.002-0.21], P=0.055 (N=377) but this was slightly stronger with the bioavailable testosterone GRS for all in males and females together: $\beta$=0.12 [95%CI:0.023-0.21], P=0.015 (N=529). There was also a weak link between our SHBG GRS, 'male SHBG cluster' [3] and IPF disease prevalence amongst males: OR=1.010 [95%CI: 1.002, 1.018], P=0.011.

### Supplementary Tables

| Disease | Number of cases, N (%) | OR | P | 95% CI | | Number of cases, N (%) | OR | P | 95% CI | |
|---|---|---|---|---|---|---|---|---|---|---|
| | MALES | | | | | FEMALES | | | | |
| All | 169,380 | | | | | 198,063 | | | | |
| IPF (J84.1) | 718 (0.41) | 0.63 | 2.2x10$^{-38}$ | 0.58 | 0.67 | 415 (0.20) | 0.69 | 8.3x10$^{-16}$ | 0.63 | 0.75 |
| Interstitial Lung Disease | 817 (0.47) | 0.64 | 1.9x10$^{-39}$ | 0.60 | 0.68 | 536 (0.26) | 0.74 | 2.1x10$^{-12}$ | 0.68 | 0.81 |
| Type II diabetes | 7,963 (4.55) | 0.91 | 2.3x10$^{-13}$ | 0.89 | 0.94 | 4,181 (2.00) | 0.91 | 1.7x10$^{-8}$ | 0.88 | 0.94 |
| Cancer incident | 9,485 (5.42) | 1.04 | 5.6x10$^{-4}$ | 1.02 | 1.06 | 8,589 (4.20) | 1.06 | 1.4x10$^{-6}$ | 1.03 | 1.08 |
| CAD | 21,331 (12.2) | 0.92 | 2.0x10$^{-28}$ | 0.90 | 0.93 | 10,119 (4.94) | 0.91 | 1.6x10$^{-18}$ | 0.89 | 0.93 |
| RA | 1,323 (0.76) | 0.87 | 9.3x10$^{-7}$ | 0.82 | 0.92 | 2,966 (1.45) | 0.93 | 2.9x10$^{-4}$ | 0.90 | 0.97 |
| GORD | 15,182 (8.68) | 0.96 | 9.8x10$^{-7}$ | 0.94 | 0.97 | 17,472 (8.53) | 0.94 | 5.7x10$^{-12}$ | 0.93 | 0.96 |
| Hyper-thyroidism | 543 (0.31) | 0.99 | 0.77 | 0.90 | 1.08 | 2,413 (1.81) | 0.94 | 3.2x10$^{-3}$ | 0.90 | 0.98 |
| Hypo-thyroidism | 2,863 (1.64) | 0.92 | 3.6x10$^{-5}$ | 0.89 | 0.96 | 15,915 (7.77) | 0.94 | 4.5x10$^{-14}$ | 0.92 | 0.95 |
| COPD | 6,565 (3.75) | 0.85 | 3.4x10$^{-35}$ | 0.83 | 0.87 | 5,330 (2.60) | 0.86 | 7.8x10$^{-24}$ | 0.84 | 0.89 |
| Anaemia | 923 (0.53) | 0.94 | 0.095 | 0.88 | 1.01 | 3,110 (1.52) | 0.996 | 0.83 | 0.96 | 1.03 |
| Participant dead | 8,151 (4.66) | 0.89 | 2.9x10$^{-24}$ | 0.86 | 0.91 | 5,192 (2.54) | 0.97 | 0.033 | 0.94 | 1.00 |

**Table S1: Associations of disease prevalence with z-adjusted $\log_e$ T/S amongst male and female participants of white European ethnicity**, with no adjustments for confounders other than age.

| Trait | Sample size | $\beta$ coeff | P | 95% CI | | Sample size | $\beta$ coeff | P | 95% CI | |
|---|---|---|---|---|---|---|---|---|---|---|
| Sex | 367,443 | -0.024 | <1.0x10$^{-314}$ | -0.024 | -0.023 | | | | | |
| | MALES | | | | | FEMALES | | | | |
| Age | 169,380 | -0.026 | <1.0x10$^{-314}$ | -0.027 | -0.025 | 198,063 | -0.021 | <1.0x10$^{-314}$ | -0.022 | -0.0208 |
| Age (>55) | 107,464 | -0.028 | <1.0x10$^{-314}$ | -0.030 | -0.027 | 121,709 | -0.028 | <1.0x10$^{-314}$ | -0.029 | -0.027 |
| Ever smoker | 166,664 | -0.056 | 2.5x10$^{-31}$ | -0.065 | -0.046 | 195,830 | -0.037 | 1.1x10$^{-16}$ | -0.046 | -0.028 |
| Former smoker | 166,664 | -0.035 | 1.7x10$^{-11}$ | -0.044 | -0.025 | 195,830 | -0.018 | 1.5x10$^{-4}$ | -0.028 | -0.009 |
| Current smoker | 166,664 | -0.13 | 1.3x10$^{-57}$ | -0.14 | -0.11 | 195,830 | -0.11 | 1.2x10$^{-41}$ | -0.13 | -0.09 |
| Pack Year Tens | 138,231 | -0.022 | 1.2x10$^{-61}$ | -0.024 | -0.019 | 168,128 | -0.023 | 1.1x10$^{-45}$ | -0.026 | -0.020 |
| BMI | 168,773 | -0.0063 | 4.1x10$^{-30}$ | -0.0074 | -0.0052 | 197,238 | -0.0064 | 1.6x10$^{-51}$ | -0.0073 | -0.0056 |
| Social deprivation | 169,168 | -0.0065 | 7.4x10$^{-17}$ | -0.0080 | -0.0049 | 197,827 | -0.0063 | 3.2x10$^{-17}$ | -0.0078 | -0.0048 |
| Fasting time | 167,377 | -0.0059 | 2.6x10$^{-10}$ | -0.0077 | -0.0041 | 198,058 | -0.0053 | 4.9x10$^{-8}$ | -0.0072 | -0.0034 |
| WBC | 164,892 | -0.017 | 1.5x10$^{-51}$ | -0.019 | -0.015 | 192,844 | -0.022 | 1.5x10$^{-84}$ | -0.025 | -0.020 |
| WBC as dependent variable | 164,892 | -0.082 | 1.5x10$^{-51}$ | -0.092 | -0.071 | 192,844 | -0.088 | 1.5x10$^{-84}$ | -0.097 | -0.079 |

**Table S2: Observational associations of key traits, in particular those known to be associated with sex hormone levels, with z-adjusted $\log_e$ T/S amongst male and female participants of white European ethnicity**, with no adjustments for confounders other than age.

| Hormone-related condition | Number of cases, % (N) | Sample size | OR for IPF | 95% CI | | P |
|---|---|---|---|---|---|---|
| Hysterectomy % (N) | 24% (30,432) | 82,058 | 1.25 | 1.002 | 1.55 | 0.048 |
| Oophorectomy % (N) | 11% (12,995) | 80,577 | 1.21 | 0.898 | 1.014 | 0.21 |
| Ever had HRT % (N) | 55% (68,383) | 81,815 | 1.46 | 1.19 | 1.80 | 0.00034 |
| Had HRT pre-2002 % (N) | 49% (55,442) | 73,724 | 1.44 | 1.15 | 1.80 | 0.0013 |
| Had HRT post-2002 % (N) | 5.7% (3,436) | 41,869 | 1.08 | 0.55 | 2.13 | 0.83 |
| Premature ovarian failure % (N) | 1.3% (1,296) | 66,914 | 3.24 | 1.85 | 5.68 | 4.2x10$^{-5}$ |
| Early menopause age <45 % (N) | 9.8% (6,563) | 41,822 | 1.80 | 1.22 | 2.65 | 3.0x10$^{-3}$ |

**Table S3: Associations of sex hormone related conditions and IPF prevalence in female unrelated individuals of European ancestry in UK Biobank for IPF cases and age-matched controls in the ratio 1:200.** Odds ratios show the extent to which IPF is associated with each trait (odds ratios and p values are adjusted for age, ever smoking, BMI and social deprivation). The number of IPF cases included in each analysis varies due to missing data for the variables involved.

| **Association N=204,736 women (F) and 174,972 men (M)** | | **IPF** | **Controls** | **IPF cases with data** | **OR** | **95% CI** | | **P** |
|---|---|---|---|---|---|---|---|---|
| Total testosterone T (F) | Concentration (SD), nmol/L | 1.13 (0.78) | 1.11 (0.62) | 247 | 1.08 | 0.92 | 1.27 | 0.34 |
| | Prevalent IPF excluded | 1.14 (0.78) | | 206 | 1.10 | 0.94 | 1.29 | 0.24 |
| | Incident IPF excluded | 1.08 (0.80) | | 41 | 0.92 | 0.54 | 1.59 | 0.78 |
| | Inverse normalised values | -0.039 | 0 | 247 | 1.02 | 0.90 | 1.15 | 0.78 |
| Total testosterone T (M) | Concentration (SD), nmol/L | 12.08 (4.06) | 11.99 (3.72) | 651 | 1.03 | 1.01 | 1.05 | 0.0018 |
| | Prevalent IPF excluded | 12.14 (4.02) | | 556 | 1.04 | 1.02 | 1.06 | 4.2x10$^{-4}$ |
| | Incident IPF excluded | 11.7 (4.28) | 0 | 95 | 0.99 | 0.94 | 1.05 | 0.76 |
| | Inverse normalised values | 0.0042 | | 651 | 1.12 | 1.03 | 1.21 | 0.0058 |
| Bioavailable T (F) | Concentration (SD), nmol/L | 0.37 (0.35) | 0.36 (0.25) | 229 | 1.10 | 0.70 | 1.72 | 0.69 |
| | Prevalent IPF excluded | 0.38 (0.37) | | 192 | 1.14 | 0.72 | 1.82 | 0.58 |
| | Incident IPF excluded | 0.36 (0.27) | | 37 | 0.82 | 0.20 | 3.39 | 0.79 |
| | Inverse normalised values | -0.008 | 0 | 229 | 0.95 | 0.83 | 1.08 | 0.42 |
| Bioavailable T % (F) | Percentage of total T | 32.1 (11.3) | 31.8 (10.5) | 229 | 0.99 | 0.97 | 1.00 | 0.04 |
| | Prevalent IPF excluded | 31.9 (11.4) | | 192 | 0.98 | 0.97 | 1.00 | 0.04 |
| | Incident IPF excluded | 33.0 (11.1) | | 37 | 0.99 | 0.96 | 1.03 | 0.70 |
| | Inverse normalised values | 0.018 | 0 | 229 | 0.85 | 0.73 | 0.97 | 0.02 |
| Bioavailable T (M) | Concentration (SD), nmol/L | 4.58 (1.34) | 5.23 (1.56) | 599 | 0.89 | 0.84 | 0.95 | 3.7x10$^{-4}$ |
| | Prevalent IPF excluded | 4.58 (1.27) | | 511 | 0.90 | 0.84 | 0.97 | 0.0032 |
| | Incident IPF excluded | 4.58 (1.70) | | 88 | 0.83 | 0.71 | 0.98 | 0.031 |
| | Inverse normalised values | -0.45 | 0 | 599 | 0.84 | 0.77 | 0.92 | 1.3x10$^{-4}$ |
| Bioavailable T % (M) | Percentage of total T | 39.7 (9.87) | 45.1 (10.4) | 599 | 0.96 | 0.955 | 0.973 | 2.7x10$^{-15}$ |
| | Prevalent IPF excluded | 39.6 (9.64) | | 511 | 0.96 | 0.95 | 0.97 | 6.3x10$^{-14}$ |
| | Incident IPF excluded | 40.6 (11.1) | | 88 | 0.97 | 0.95 | 0.99 | 0.012 |
| | Inverse normalised values | -0.53 | 0 | 599 | **0.69** | **0.63** | **0.76** | **5.6x10$^{-16}$** |
| Free T (F) | Concentration (SD), pmol/L | 15.7 (15.4) | 14.6 (10.3) | 229 | 57 | 0.003 | 106 | 0.42 |
| | Prevalent IPF excluded | 15.8 (16.1) | | 192 | 144 | 0.007 | 106 | 0.32 |
| | Incident IPF excluded | 15.0 (0.8) | | 37 | 0.026 | - | - | |
| | Inverse normalised values | 0.035 | 0 | 229 | 0.98 | 0.85 | 1.12 | 0.75 |
| Free T % (F) | Percentage of total T | 1.34 (0.47) | 1.30 (0.43) | 229 | 0.80 | 0.57 | 1.13 | 0.21 |
| | Prevalent IPF excluded | 1.38 (0.47) | | 192 | 0.78 | 0.54 | 1.13 | 0.19 |
| | Incident IPF excluded | 1.38 (0.43) | | 37 | 0.94 | 0.41 | 2.15 | 0.89 |
| | Inverse normalised values | 0.018 | | 229 | 0.85 | 0.73 | 0.97 | 0.02 |
| Free T (M) | Concentration (SD), pmol/L | 192 (56) | 211 (62) | 599 | 0.35 | 0.078 | 1.60 | 0.18 |
| | Prevalent IPF excluded | 192 (53) | | 511 | 0.42 | 0.08 | 2.13 | 0.29 |
| | Incident IPF excluded | 194 (71) | | 88 | 0.14 | 0.003 | 7.47 | 0.34 |
| | Inverse normalised values | -0.33 | 0 | 599 | 0.94 | 0.86 | 1.02 | 0.13 |
| Free T % (M) | Percentage of total T | 1.67 (0.41) | 1.82 (0.41) | 599 | 0.55 | 0.44 | 0.69 | 1.6x10$^{-7}$ |
| | Prevalent IPF excluded | 1.66 (0.40) | | 511 | 0.52 | 0.41 | 0.66 | 1.4x10$^{-7}$ |
| | Incident IPF excluded | 1.72 (0.46) | | 88 | 0.75 | 0.43 | 1.34 | 0.34 |
| | Inverse normalised values | -0.53 | | 599 | 0.69 | 0.63 | 0.76 | 5.6x10$^{-16}$ |
| Sex Hormone Binding Globulin (SHBG) (F) | Concentration (SD), nmol/L | 60.4 (30.9) | 62.4 (31.0) | 364 | 1.003 | 0.999 | 1.007 | 0.10 |
| | Prevalent IPF excluded | 61.4 (31.8) | | 281 | 1.005 | 1.001 | 1.009 | 0.018 |
| | Incident IPF excluded | 56.7 (27.2) | | 65 | 0.99 | 0.98 | 1.004 | 0.026 |
| | Inverse normalised values | -0.082 (1.06) | 0 | 364 | 1.083 | 0.96 | 1.22 | 0.18 |
| Sex Hormone Binding Globulin (SHBG) (M) | Concentration (SD), nmol/L | 47.7 (20.4) | 39.9 (16.8) | 608 | 1.015 | 1.011 | 1.019 | 6.9x10$^{-13}$ |
| | Prevalent IPF excluded | 47.9 (19.9) | | 518 | 1.015 | 1.011 | 1.019 | 4.5x10$^{-12}$ |
| | Incident IPF excluded | 46.5 (23.6) | | 90 | 1.011 | 1.005 | 1.022 | 0.04 |
| | Inverse normalised values | 0.43 (1.02) | 0 | 608 | **1.35** | **1.23** | **1.47** | **2.6x10$^{-11}$** |
| Albumin (F) | Concentration (SD), g/L | 43.7 (2.7) | 45.0 (2.6) | 349 | **0.86** | **0.82** | **0.89** | **4.2x10$^{-14}$** |
| | Prevalent IPF excluded | 43.8 (2.8) | | 284 | 0.87 | 0.83 | 0.91 | 5.7x10$^{-10}$ |
| | Incident IPF excluded | 43.4 (2.3) | | 65 | 0.81 | 0.74 | 0.89 | 4.9x10$^{-6}$ |
| | Inverse normalised values | -0.48 (1.03) | 0 | 349 | **0.66** | **0.60** | **0.74** | **1.3x10$^{-13}$** |
| Albumin (M) | Concentration (SD), g/L | 43.8 (2.9) | 45.5 (2.6) | 614 | **0.84** | **0.82** | **0.86** | **3.9x10$^{-34}$** |
| | Prevalent IPF excluded | 43.9 (2.9) | | 523 | 0.85 | 0.83 | 0.88 | 1.3x10$^{-24}$ |
| | Incident IPF excluded | 43.3 (3.0) | | 91 | 0.80 | 0.75 | 0.85 | 2.0x10$^{-12}$ |
| | Inverse normalised values | -0.66 (1.05) | 0 | 614 | **0.62** | **0.57** | **0.67** | **7.7x10$^{-31}$** |

**Table S4: Associations between IPF prevalence and recruitment measured levels of sex hormone levels amongst male unrelated individuals of European ancestry in UK Biobank derived using logistic regression analyses.** Odds ratios show the extent to which the hormone concentrations predicted IPF outcomes (odds ratios and p values are adjusted for age, ever smoking, BMI, deprivation and fasting time). The proportion of IPF cases included in each analysis varies due to missing data for the variables involved. Bioavailable and free testosterone associations are shown for absolute values and percentages of total testosterone.

| IPF cases with HRT | HRT variable | Pre, post 2002 or all | IPF cases with data | $\beta$ coef | 95% CI | | P |
|---|---|---|---|---|---|---|---|
| Age of IPF diagnosis | HRT start age | All HRT | 204 | 0.44 | 0.28 | 0.61 | $2.3 \times 10^{-7}$ |
| | **HRT start age** | **HRT Pre 2002** | **191** | **0.50** | **0.34** | **0.65** | $\mathbf{2.3 \times 10^{-9}}$ |
| | HRT start age | HRT Post 2002 | 8 | 1.58 | 0.81 | 2.36 | 0.0075 |
| | **HRT end age (ongoing included)** | **All HRT** | **197** | **0.41** | **0.27** | **0.54** | $\mathbf{3.4 \times 10^{-8}}$ |
| | HRT end age (ongoing included) | HRT Pre 2002 | 175 | 0.35 | 0.21 | 0.48 | $1.9 \times 10^{-6}$ |
| | HRT end age (ongoing included) | HRT Post 2002 | 8 | 1.77 | 0.81 | 2.72 | 0.0098 |
| | HRT treatment duration | All HRT | 136 | 0.11 | -0.09 | 0.30 | 0.27 |
| Age of death | HRT start age | All HRT | 62 | 0.27 | 0.05 | 0.49 | 0.017 |
| | **HRT start age** | **HRT Pre 2002** | **58** | **0.31** | **0.13** | **0.49** | **0.0013** |
| | HRT end age (ongoing included) | All HRT | 57 | 0.18 | 0.003 | 0.36 | 0.047 |
| | HRT end age (ongoing included) | HRT Pre 2002 | 51 | 0.15 | -0.015 | 0.31 | 0.074 |
| | HRT treatment duration | All HRT | 41 | 0.03 | -0.26 | 0.32 | 0.84 |

**Table S5: Associations between age of IPF diagnosis or death and HRT parameters in female unrelated individuals of European ancestry in UK Biobank derived using linear regression analyses.** Beta coefficients show the extent to which the exposures predicted IPF outcomes (analyses are adjusted for ever smoking, BMI and deprivation but not age, since age of diagnosis and age of registration are very highly correlated for this disease due to poor survival ($\boldsymbol{\beta}$=1.10, $p<10^{-104}$)). The proportion of IPF cases included in each analysis varies due to missing data for the variables involved.

### **Tables S6a & S6b.** Observational associations of leukocyte telomere length with sex hormone levels in females and males

| **Female Participants N = 204,736** | IPF N=415 | IPF cases with data | Controls N=204,321 | Sample size | $\beta$ coeff | 95% CI | | P |
|---|---|---|---|---|---|---|---|---|
| Hysterectomy % (N) | 28% (116) | 415 | 19% (38,291) | 69,288 | 0.031 | -0.0063 | 0.067 | 0.10 |
| Oophorectomy % (N) | 12% (49) | 402 | 8.1% (15,478) | 69,287 | 0.058 | 0.012 | 0.103 | 0.014 |
| Ever had HRT % (N) | 62% (256) | 413 | 39% (79,967) | 69,109 | -0.034 | -0.049 | -0.019 | $1.3 \times 10^{-5}$ |
| Had HRT pre-2002 % (N) | 49% (204) | 361 | 29.5% (60,168) | 65,885 | -0.036 | -0.052 | -0.020 | $1.3 \times 10^{-5}$ |
| Had HRT post-2002 % (N) | 2.2% (9) | 166 | 4.2% (8,482) | 46,964 | -0.011 | -0.054 | 0.032 | 0.61 |
| Premature ovarian failure % (N) | 3.1% (13) | 309 | 0.97% (1,982) | 69,288 | -0.059 | -0.113 | -0.004 | 0.036 |
| Early menopause age <45 % (N) | 8.2% (34) | 180 | 4.5% (9,284) | 54,475 | -0.087 | -0.112 | -0.061 | $2.9 \times 10^{-11}$ |

**Table S6a: Associations of Z-adjusted LTL Log with sex hormone related conditions in female unrelated individuals of European ancestry over the age of 55 at registration in UK Biobank**. Beta coefficients show the extent to which the traits are associated with LTL (analyses were adjusted for age, ever smoking, BMI and social deprivation).

| Trait | Sample size | $\beta$ coeff | P | 95% CI | | Sample size | $\beta$ coeff | P | 95% CI | |
|---|---|---|---|---|---|---|---|---|---|---|
| | MALES | | | | | FEMALES | | | | |
| SHBG | 145,285 | -0.0028 | $3.2 \times 10^{-65}$ | -0.0031 | -0.0025 | 167,166 | -0.000084 | $3.2 \times 10^{-9}$ | -0.00066 | -0.00033 |
| SHBG (inverse normalised) | 145,285 | -0.047 | $3.3 \times 10^{-62}$ | -0.052 | -0.041 | 167,166 | -0.018 | $1.1 \times 10^{-11}$ | -0.023 | -0.013 |
| Total testosterone (T) | 156,961 | -0.0084 | $1.0 \times 10^{-33}$ | -0.0097 | -0.0070 | 155,121 | -0.012 | $2.0 \times 10^{-3}$ | -0.020 | -0.0045 |
| Total T (inv. norm.) | 156,961 | -0.032 | $1.4 \times 10^{-34}$ | -0.037 | -0.027 | 155,121 | -0.0070 | $5.5 \times 10^{-3}$ | -0.012 | -0.002 |
| Bioavailable T % | 144,342 | 0.0043 | $1.6 \times 10^{-54}$ | 0.0037 | 0.0048 | 140,309 | 0.0018 | $5.8 \times 10^{-11}$ | 0.00125 | 0.00233 |
| Bioavailable T % (inv. norm.) | 144,342 | 0.045 | $2.3 \times 10^{-57}$ | 0.040 | 0.051 | 140,309 | 0.0188 | $5.8 \times 10^{-11}$ | 0.013 | 0.024 |
| Free T % | 144,342 | 0.109 | $3.0 \times 10^{-56}$ | 0.095 | 0.122 | 140,309 | 0.046 | $1.9 \times 10^{-11}$ | 0.033 | 0.060 |
| Free T % (inv. norm.) | 144,342 | 0.045 | $2.3 \times 10^{-57}$ | 0.040 | 0.051 | 140,309 | 0.0188 | $5.8 \times 10^{-11}$ | 0.013 | 0.024 |
| Albumin | 146,405 | 0.0009 | 0.36 | -0.0011 | 0.0029 | 169,644 | 0.00040 | 0.67 | -0.0014 | 0.0022 |
| Albumin (inv. norm.) | 146,405 | 0.0021 | 0.43 | -0.0031 | 0.0072 | 169,644 | 0.00096 | 0.69 | -0.0038 | 0.0057 |

**Table S6b: Associations of Z-adjusted LTL Log with sex hormone levels amongst male and female unrelated individuals of European ancestry in UK Biobank.** Beta coefficients show the extent to which the hormone levels are associated with LTL (analyses were adjusted for age, ever smoking, BMI, social deprivation and fasting time).

**Tables S7a & S7b.** Cox proportional regression analysis of time from registration to IPF diagnosis amongst all and time to IPF death amongst IPF cases, with respect to LTL and age of natural menopause for females in UK Biobank

| Trait | Sample size | HR | P | 95% CI | |
|---|---|---|---|---|---|
| (i) LTL log adjusted only for age | 197,952 | 0.69 | $6.4 \times 10^{-18}$ | 0.63 | 0.75 |
| Age | | 1.12 | $5.2 \times 10^{-29}$ | 1.10 | 1.14 |
| (ii) LTL log adjusted for age and menopause age | 87,791 | 0.64 | $9.0 \times 10^{-10}$ | 0.55 | 0.74 |
| Age | | 1.10 | $8.0 \times 10^{-10}$ | 1.07 | 1.14 |
| Age of natural menopause | | 0.97 | 0.028 | 0.94 | 0.996 |
| (iii) LTL log with multiple covariates | 86,584 | 0.64 | $1.3 \times 10^{-9}$ | 0.63 | 0.76 |
| Age | | 1.10 | $5.0 \times 10^{-9}$ | 1.09 | 1.14 |
| Age of natural menopause | | 0.97 | 0.044 | 0.94 | 0.999 |
| Ever smoker | | 2.56 | $5.3 \times 10^{-8}$ | 1.79 | 3.09 |
| Body mass index | | 1.03 | 0.019 | 1.02 | 1.07 |
| Social deprivation | | 1.03 | 0.27 | 1.02 | 1.11 |

**Table S7a:** Cox proportional hazard analyses of time from registration to diagnosis of IPF with respect to (i) LTL adjusted for age only (amongst all females), (ii) LTL adjusted for age and age of natural menopause (amongst those who had natural menopause) and (iii) LTL adjusted for age and age of natural menopause with additional adjustment for sex hormone level confounders (amongst those who had natural menopause). Testing showed no evidence that the proportional hazards assumptions were violated.

| Trait | Sample size | HR | P | 95% CI | |
|---|---|---|---|---|---|
| (i) LTL log adjusted only for age | 193,056 | 0.64 | $2.0 \times 10^{-17}$ | 0.58 | 0.71 |
| Age | | 1.12 | $6.9 \times 10^{-13}$ | 1.09 | 1.16 |
| (ii) LTL log adjusted for age and menopause age | 85,307 | 0.57 | $1.5 \times 10^{-8}$ | 0.47 | 0.69 |
| Age | | 1.10 | $6.8 \times 10^{-5}$ | 1.05 | 1.16 |
| Age of natural menopause | | 0.94 | $7.6 \times 10^{-3}$ | 0.90 | 0.98 |
| (iii) LTL log with multiple covariates | 84,174 | 0.57 | $6.4 \times 10^{-9}$ | 0.47 | 0.69 |
| Age | | 1.10 | $9.3 \times 10^{-5}$ | 1.05 | 1.16 |
| Age of natural menopause | | 0.95 | 0.025 | 0.90 | 0.99 |
| Ever smoker | | 2.09 | $4.3 \times 10^{-3}$ | 1.26 | 3.48 |
| Body mass index | | 1.07 | $1.1 \times 10^{-3}$ | 1.03 | 1.11 |
| Social deprivation | | 0.996 | 0.94 | 0.92 | 1.08 |

**Table 7b:** Cox proportional hazard analyses of time from registration to death with IPF with respect to (i) LTL adjusted for age only (amongst all females), (ii) LTL adjusted for age and age of natural menopause (amongst those who had natural menopause) and (iii) LTL adjusted for age and age of natural menopause with additional adjustment for sex hormone level confounders (amongst those who had natural menopause). Testing showed no evidence that the proportional hazards assumptions were violated.

| Trait | Sample size | HR | P | 95% CI | |
|---|---|---|---|---|---|
| (i) LTL log adjusted only for age | 259 | 0.82 | 0.044 | 0.67 | 0.99 |
| Age | | 1.02 | 0.33 | 0.98 | 1.07 |
| (ii) LTL log adjusted for age and HRT use pre 2002 | 248 | 0.77 | 0.022 | 0.61 | 0.96 |
| Age | | 1.03 | 0.27 | 0.98 | 1.08 |
| Received HRT (pre-2002) | | 0.59 | 0.033 | 0.37 | 0.96 |
| (iii) LTL log with multiple covariates | 241 | 0.75 | 0.016 | 0.59 | 0.95 |
| Age | | 1.03 | 0.23 | 0.98 | 1.08 |
| Received HRT (pre-2002) | | 0.60 | 0.041 | 0.37 | 0.98 |
| Ever smoker | | 1.05 | 0.83 | 0.65 | 1.72 |
| Body mass index | | 1.02 | 0.36 | 0.98 | 1.06 |
| Social deprivation | | 1.01 | 0.78 | 0.94 | 1.09 |

**Table S8: Cox proportional regression analysis of time from diagnosis to death amongst IPF cases with respect to LTL and prior receipt of HRT for post-menopausal females in UK Biobank**
Cox proportional hazard analyses of time from diagnosis of IPF to death amongst post-menopausal females, with respect to (i) LTL adjusted for age only, (ii) LTL adjusted for age and whether they received HRT and (iii) LTL adjusted for age and receipt of HRT with additional adjustment for sex hormone level confounders. Testing showed no evidence that the proportional hazards assumptions were violated.

**Tables S9a-i.** Cox proportional regression models for time from registration to incident cases of IPF diagnosis amongst all and time to IPF death amongst IPF cases, with respect to LTL and bioavailable testosterone, free testosterone and SHBG for males in UK Biobank

| Trait | Sample size | HR | P | 95% CI | |
|---|---|---|---|---|---|
| (i) LTL log adjusted only for age | 169,146 | 0.61 | $2.4x10^{-39}$ | 0.57 | 0.66 |
| Age | | 1.13 | $3.2x10^{-53}$ | 1.11 | 1.15 |
| (ii) LTL log adjusted for age and bioavailable T % | 147,179 | 0.64 | $2.1x10^{-25}$ | 0.59 | 0.70 |
| Age | | 1.11 | $6.3x10^{-35}$ | 1.10 | 1.13 |
| **Bioavailable testosterone %** | | **0.97** | **$8.7x10^{-10}$** | **0.96** | **0.98** |
| (iii) LTL log with multiple covariates | 144,146 | 0.65 | $3.8x10^{-23}$ | 0.60 | 0.71 |
| Age | | 1.11 | $6.8x10^{-30}$ | 1.09 | 1.13 |
| **Bioavailable testosterone %** | | **0.96** | **$6.4x10^{-12}$** | **0.95** | **0.97** |
| Ever smoker | | 2.11 | $5.7x10^{-12}$ | 1.70 | 2.60 |
| Body mass index | | 1.05 | $1.1x10^{-6}$ | 1.03 | 1.07 |
| Social deprivation | | 1.05 | $1.1x10^{-3}$ | 1.02 | 1.08 |
| Fasting time | | 1.00 | 0.96 | 0.96 | 1.04 |
| (iv) LTL log adjusted for age and bioavailable T | 147,179 | 0.64 | $7.8x10^{-27}$ | 0.59 | 0.69 |
| Age | | 1.12 | $8.5x10^{-38}$ | 1.10 | 1.14 |
| **Bioavailable testosterone concentration** | | **0.88** | **$3.6x10^{-4}$** | **0.82** | **0.94** |
| (v) LTL log with multiple covariates | 144,146 | 0.64 | $4.9x10^{-25}$ | 0.59 | 0.70 |
| Age | | 1.12 | $1.2x10^{-34}$ | 1.10 | 1.14 |
| **Bioavailable testosterone concentration** | | **0.91** | **$9.3x10^{-3}$** | **0.85** | **0.98** |
| Ever smoker | | 2.08 | $1.4x10^{-11}$ | 1.68 | 2.57 |
| Body mass index | | 1.03 | 0.011 | 1.01 | 1.05 |
| Social deprivation | | 1.05 | $3.6x10^{-4}$ | 1.02 | 1.08 |
| Fasting time | | 1.004 | 0.80 | 0.97 | 1.04 |

**Table S9a: Cox proportional hazard analyses of time from registration to diagnosis of IPF** with respect to (i) LTL adjusted for age only (amongst all males), (ii) LTL adjusted for age and bioavailable testosterone %, (iii) LTL adjusted for age and bioavailable testosterone % with additional adjustment for sex hormone level confounders (iv) LTL adjusted for age and bioavailable testosterone concentration and (v) LTL adjusted for age and bioavailable testosterone concentration with additional adjustment for sex hormone level confounders. Testing showed no evidence that the proportional hazards assumptions were violated.

| Trait | Sample size | HR | P | 95% CI | |
|---|---|---|---|---|---|
| (i) LTL log adjusted only for age | 169,146 | 0.61 | $2.4 \times 10^{-39}$ | 0.57 | 0.66 |
| Age | | 1.13 | $3.2 \times 10^{-53}$ | 1.11 | 1.15 |
| (ii) LTL log adjusted for age and free T % | 147,179 | 0.64 | $5.2 \times 10^{-26}$ | 0.59 | 0.70 |
| Age | | 1.12 | $3.0 \times 10^{-39}$ | 1.10 | 1.14 |
| **Free testosterone %** | | **0.62** | **$1.1 \times 10^{-4}$** | **0.49** | **0.79** |
| (iii) LTL log with multiple covariates | 144,146 | 0.65 | $9.5 \times 10^{-24}$ | 0.60 | 0.71 |
| Age | | 1.11 | $1.1 \times 10^{-33}$ | 1.09 | 1.13 |
| **Free testosterone %** | | **0.54** | **$2.1 \times 10^{-6}$** | **0.42** | **0.70** |
| Ever smoker | | 2.11 | $5.0 \times 10^{-12}$ | 1.71 | 2.61 |
| Body mass index | | 1.05 | $1.5 \times 10^{-5}$ | 1.03 | 1.07 |
| Social deprivation | | 1.05 | $6.2 \times 10^{-4}$ | 1.02 | 1.08 |
| Fasting time | | 1.00 | 0.96 | 0.96 | 1.04 |
| (ii) LTL log adjusted for age and free T concentration | 147,179 | 0.64 | $6.3 \times 10^{-27}$ | 0.59 | 0.69 |
| Age | | 1.13 | $1.5 \times 10^{-41}$ | 1.11 | 1.14 |
| Free testosterone concentration | | 0.23 | 0.093 | 0.04 | 1.28 |
| (iii) LTL log with multiple covariates | 144,146 | 0.64 | $4.4 \times 10^{-25}$ | 0.59 | 0.70 |
| Age | | 1.12 | $8.4 \times 10^{-38}$ | 1.10 | 1.14 |
| Free testosterone concentration | | 0.50 | 0.42 | 0.09 | 2.71 |
| Ever smoker | | 2.09 | $8.8 \times 10^{-12}$ | 1.69 | 2.59 |
| Body mass index | | 1.03 | 0.0048 | 1.01 | 1.05 |
| Social deprivation | | 1.05 | $3.3 \times 10^{-4}$ | 1.02 | 1.08 |
| Fasting time | | 1.003 | 0.89 | 0.97 | 1.04 |

**Table S9b: Cox proportional hazard analyses of time from registration to diagnosis of IPF** with respect to (i) LTL adjusted for age only (amongst all males), (ii) LTL adjusted for age and free testosterone % and (iii) LTL adjusted for age and free testosterone % with additional adjustment for sex hormone level confounders, (iv) LTL adjusted for age and free testosterone concentration and (v) LTL adjusted for age and free testosterone concentration with additional adjustment for sex hormone level confounders. Testing showed no evidence that the proportional hazards assumptions were violated.

| Trait | Sample size | HR | P | 95% CI | |
|---|---|---|---|---|---|
| (i) LTL log adjusted only for age | 169,146 | 0.61 | $2.4 \times 10^{-39}$ | 0.57 | 0.66 |
| Age | | 1.13 | $3.2 \times 10^{-53}$ | 1.11 | 1.15 |
| (ii) LTL log adjusted for age and SHBG | 148,138 | 0.64 | $1.4 \times 10^{-26}$ | 0.59 | 0.69 |
| Age | | 1.12 | $9.8 \times 10^{-40}$ | 1.10 | 1.14 |
| **SHBG** | | **1.012** | **$1.2 \times 10^{-7}$** | **1.007** | **1.016** |
| (iii) LTL log with multiple covariates | 145,086 | 0.65 | $2.6 \times 10^{-24}$ | 0.59 | 0.70 |
| Age | | 1.11 | $1.5 \times 10^{-34}$ | 1.09 | 1.13 |
| **SHBG** | | **1.014** | **$5.9 \times 10^{-10}$** | **1.010** | **1.019** |
| Ever smoker | | 2.13 | $2.3 \times 10^{-12}$ | 1.72 | 2.63 |
| Body mass index | | 1.05 | $5.8 \times 10^{-6}$ | 1.03 | 1.07 |
| Social deprivation | | 1.05 | $1.1 \times 10^{-3}$ | 1.02 | 1.08 |
| Fasting time | | 1.00 | 0.85 | 0.96 | 1.03 |

**Table S9c: Cox proportional hazard analyses of time from registration to diagnosis** of IPF with respect to (i) LTL adjusted for age only (amongst all males), (ii) LTL adjusted for age and SHBG and (iii) LTL adjusted for age and SHBG with additional adjustment for sex hormone level confounders. Testing showed no evidence that the proportional hazards assumptions were violated.

| Trait | Sample size | HR | P | 95% CI | |
|---|---|---|---|---|---|
| (i) LTL log adjusted only for age | 161,721 | 0.59 | $5.1 \times 10^{-28}$ | 0.54 | 0.65 |
| Age | | 1.17 | $7.8 \times 10^{-38}$ | 1.14 | 1.20 |
| (ii) LTL log adjusted for age and bioavailable T | 140,786 | 0.62 | $2.6 \times 10^{-18}$ | 0.55 | 0.69 |
| Age | | 1.15 | $5.6 \times 10^{-26}$ | 1.12 | 1.18 |
| **Bioavailable testosterone %** | | **0.96** | **$1.4 \times 10^{-8}$** | **0.948** | **0.974** |
| (iii) LTL log with multiple covariates | 137,996 | 0.63 | $4.9 \times 10^{-16}$ | 0.56 | 0.70 |
| Age | | 1.14 | $3.3 \times 10^{-23}$ | 1.11 | 1.17 |
| **Bioavailable testosterone %** | | **0.96** | **$1.2 \times 10^{-9}$** | **0.94** | **0.97** |
| Ever smoker | | 2.35 | $1.6 \times 10^{-8}$ | 1.75 | 3.16 |
| Body mass index | | 1.04 | $4.4 \times 10^{-3}$ | 1.01 | 1.07 |
| Social deprivation | | 1.07 | $2.5 \times 10^{-4}$ | 1.03 | 1.11 |
| Fasting time | | 1.03 | 0.17 | 0.99 | 1.08 |
| (iv) LTL log adjusted for bioavailable testosterone | 140,786 | 0.61 | $1.8 \times 10^{-19}$ | 0.55 | 0.68 |
| Age | | 1.15 | $7.3 \times 10^{-27}$ | 1.12 | 1.18 |
| **Bioavailable testosterone concentration** | | **0.78** | **$2.4 \times 10^{-6}$** | **0.71** | **0.87** |
| (v) LTL log with multiple covariates | 137,996 | 0.62 | $1.5 \times 10^{-17}$ | 0.56 | 0.70 |
| Age | | 1.15 | $1.1 \times 10^{-24}$ | 1.12 | 1.18 |
| **Bioavailable testosterone concentration** | | **0.80** | **$2.9 \times 10^{-5}$** | **0.73** | **0.89** |
| Ever smoker | | 2.29 | $4.7 \times 10^{-8}$ | 1.70 | 3.08 |
| Body mass index | | 1.01 | 0.63 | 0.98 | 1.04 |
| Social deprivation | | 1.08 | $8.8 \times 10^{-5}$ | 1.04 | 1.12 |
| Fasting time | | 1.04 | 0.072 | 0.996 | 1.09 |

**Table S9d**: **Cox proportional hazard analyses of time from registration to death from IPF** with respect to (i) LTL adjusted for age only (amongst all males), (ii) LTL adjusted for age and bioavailable testosterone % and (iii) LTL adjusted for age and bioavailable testosterone % with additional adjustment for sex hormone level confounders, (iv) LTL adjusted for age and bioavailable testosterone concentration and (v) LTL adjusted for age and bioavailable testosterone concentration with additional adjustment for sex hormone level confounders. Testing showed no evidence that the proportional hazards assumptions were violated.

| Trait | Sample size | HR | P | 95% CI | |
|---|---|---|---|---|---|
| (i) LTL log adjusted only for age | 161,721 | 0.59 | $5.1 \times 10^{-28}$ | 0.54 | 0.65 |
| Age | | 1.17 | $7.8 \times 10^{-38}$ | 1.14 | 1.20 |
| (ii) LTL log adjusted for age and free T % | 140,786 | 0.62 | $7.7 \times 10^{-19}$ | 0.55 | 0.69 |
| Age | | 1.16 | $3.1 \times 10^{-29}$ | 1.13 | 1.19 |
| **Free testosterone %** | | **0.57** | **$9.1 \times 10^{-4}$** | **0.41** | **0.79** |
| (iii) LTL log with multiple covariates | 137,996 | 0.63 | $1.3 \times 10^{-16}$ | 0.56 | 0.70 |
| Age | | 1.15 | $4.0 \times 10^{-26}$ | 1.12 | 1.18 |
| **Free testosterone %** | | **0.51** | **$1.6 \times 10^{-4}$** | **0.36** | **0.72** |
| Ever smoker | | 2.35 | $1.5 \times 10^{-8}$ | 1.75 | 3.17 |
| Body mass index | | 1.03 | $2.6 \times 10^{-2}$ | 1.004 | 1.07 |
| Social deprivation | | 1.08 | $1.4 \times 10^{-4}$ | 1.04 | 1.12 |
| Fasting time | | 1.03 | 0.17 | 0.99 | 1.08 |
| (ii) LTL log adjusted for age and free T concentration | 140,786 | 0.61 | $1.7 \times 10^{-19}$ | 0.55 | 0.68 |
| Age | | 1.16 | $1.2 \times 10^{-29}$ | 1.13 | 1.19 |
| **Free testosterone concentration** | | **0.036** | **0.0073** | **0.003** | **0.408** |
| (iii) LTL log with multiple covariates | 137,996 | 0.62 | $1.5 \times 10^{-17}$ | 0.56 | 0.70 |
| Age | | 1.16 | $3.2 \times 10^{-27}$ | 1.13 | 1.19 |
| **Free testosterone concentration** | | **0.064** | **0.028** | **0.005** | **0.75** |
| Ever smoker | | 2.31 | $3.1 \times 10^{-8}$ | 1.72 | 3.11 |
| Body mass index | | 1.01 | 0.42 | 0.98 | 1.04 |
| Social deprivation | | 1.08 | $8.0 \times 10^{-5}$ | 1.04 | 1.12 |
| Fasting time | | 1.04 | 0.1 | 0.99 | 1.05 |

**Table S9e: Cox proportional hazard analyses of time from registration to death from IPF** with respect to (i) LTL adjusted for age only (amongst all males), (ii) LTL adjusted for age and free testosterone % and (iii) LTL adjusted for age and free testosterone % with additional adjustment for sex hormone level confounders, (iv) LTL adjusted for age and free testosterone concentration and (v) LTL adjusted for age and free testosterone concentration with additional adjustment for sex hormone level confounders. Testing showed no evidence that the proportional hazards assumptions were violated.

| Trait | Sample size | HR | P | 95% CI | |
|---|---|---|---|---|---|
| (i) LTL log adjusted only for age | 161,721 | 0.59 | $5.1 \times 10^{-28}$ | 0.54 | 0.65 |
| Age | | 1.17 | $7.8 \times 10^{-38}$ | 1.14 | 1.20 |
| (ii) LTL log adjusted for SHBG | 141,638 | 0.62 | $3.5 \times 10^{-19}$ | 0.55 | 0.68 |
| Age | | 1.16 | $5.0 \times 10^{-30}$ | 1.13 | 1.19 |
| **SHBG** | | **1.014** | **$2.5 \times 10^{-6}$** | **1.008** | **1.020** |
| (iii) LTL log with multiple covariates | 138,834 | 0.63 | $6.5 \times 10^{-17}$ | 0.56 | 0.70 |
| Age | | 1.15 | $5.4 \times 10^{-27}$ | 1.12 | 1.18 |
| **SHBG** | | **1.016** | **$2.5 \times 10^{-7}$** | **1.010** | **1.022** |
| Ever smoker | | 2.42 | $4.3 \times 10^{-9}$ | 1.80 | 3.26 |
| Body mass index | | 1.04 | 0.013 | 1.01 | 1.07 |
| Social deprivation | | 1.07 | $2.2 \times 10^{-4}$ | 1.03 | 1.11 |
| Fasting time | | 1.03 | 0.24 | 0.98 | 1.07 |

**Table S9f: Cox proportional hazard analyses of time from registration to death with IPF** with respect to (i) LTL adjusted for age only (amongst all males), (ii) LTL adjusted for age and SHBG and (iii) LTL adjusted for age and SHBG with additional adjustment for sex hormone level confounders. Testing showed no evidence that the proportional hazards assumptions were violated.

| Trait | Sample size | HR | P | 95% CI | |
|---|---|---|---|---|---|
| (i) LTL log adjusted only for age | 694 | 0.93 | 0.098 | 0.85 | 1.01 |
| Age | | 1.04 | $4.9 \times 10^{-4}$ | 1.02 | 1.07 |
| (ii) LTL log adjusted for age and bioavailable T % | 595 | 0.57 | $1.5 \times 10^{-8}$ | 0.47 | 0.69 |
| Age | | 1.04 | 0.0044 | 1.01 | 1.06 |
| **Bioavailable testosterone %** | | **0.98** | **0.040** | **0.97** | **1.00** |
| (iii) LTL log with multiple covariates | 578 | 0.95 | 0.30 | 0.86 | 1.05 |
| Age | | 1.04 | 0.0027 | 1.01 | 1.07 |
| Bioavailable testosterone % | | 0.99 | 0.23 | 0.98 | 1.005 |
| Ever smoker | | 1.06 | 0.70 | 0.78 | 1.44 |
| Body mass index | | 0.99 | 0.38 | 0.97 | 1.02 |
| Social deprivation | | 1.00 | 0.91 | 0.96 | 1.04 |
| Fasting time | | 1.04 | 0.10 | 0.99 | 1.08 |
| (iv) LTL log adjusted for bioavailable testosterone | 594 | 0.95 | 0.29 | 0.87 | 1.04 |
| Age | | 1.04 | 0.0031 | 1.01 | 1.07 |
| **Bioavailable testosterone concentration** | | **0.89** | **0.023** | **0.81** | **0.98** |
| (v) LTL log with multiple covariates | 578 | 0.95 | 0.29 | 0.86 | 1.04 |
| Age | | 1.04 | 0.0059 | 1.01 | 1.07 |
| **Bioavailable testosterone concentration** | | **0.89** | **0.018** | **0.80** | **0.98** |
| Ever smoker | | 1.02 | 0.89 | 0.76 | 1.38 |
| Body mass index | | 0.98 | 0.25 | 0.96 | 1.01 |
| Social deprivation | | 1.00 | 1.00 | 0.96 | 1.04 |
| Fasting time | | 1.04 | 0.065 | 0.998 | 1.09 |

**Table S9g: Cox proportional hazard analyses of time from registration to death with IPF** with respect to (i) LTL adjusted for age only (amongst all males), (ii) LTL adjusted for age and bioavailable testosterone % and (iii) LTL adjusted for age and bioavailable testosterone % with additional adjustment for sex hormone level confounders, (iv) LTL adjusted for age and bioavailable testosterone concentration and (v) LTL adjusted for age and bioavailable testosterone concentration with additional adjustment for sex hormone level confounders. Testing showed no evidence that the proportional hazards assumptions were violated.

| Trait | Sample size | HR | P | 95% CI | |
|---|---|---|---|---|---|
| (i) LTL log adjusted only for age | 694 | 0.93 | 0.098 | 0.85 | 1.01 |
| Age | | 1.04 | $4.9x10^{-4}$ | 1.02 | 1.07 |
| (ii) LTL log adjusted for age and free T % | 594 | 0.95 | 0.28 | 0.86 | 1.04 |
| Age | | 1.04 | 0.0077 | 1.02 | 1.07 |
| Free testosterone % | | 0.89 | 0.49 | 0.65 | 1.23 |
| (iii) LTL log with multiple covariates | 578 | 0.95 | 0.27 | 0.86 | 1.04 |
| Age | | 1.04 | 0.0017 | 1.02 | 1.07 |
| Free testosterone % | | 0.93 | 0.70 | 0.66 | 1.32 |
| Ever smoker | | 1.05 | 0.75 | 0.78 | 1.43 |
| Body mass index | | 0.99 | 0.40 | 0.96 | 1.02 |
| Social deprivation | | 1.00 | 0.92 | 0.96 | 1.04 |
| Fasting time | | 1.04 | 0.087 | 0.99 | 1.08 |
| (ii) LTL log adjusted for age and free T concentration | 594 | 0.95 | 0.29 | 0.87 | 1.04 |
| Age | | 1.04 | 0.0019 | 1.015 | 1.07 |
| Free testosterone concentration | | 0.14 | 0.10 | 0.014 | 1.49 |
| (iii) LTL log with multiple covariates | 578 | 0.95 | 0.28 | 0.86 | 1.04 |
| Age | | 1.04 | 0.0037 | 1.01 | 1.07 |
| Free testosterone concentration | | 0.13 | 0.097 | 0.012 | 1.44 |
| Ever smoker | | 1.03 | 0.84 | 0.76 | 1.40 |
| Body mass index | | 0.98 | 0.28 | 0.96 | 1.01 |
| Social deprivation | | 1.00 | 0.99 | 0.96 | 1.04 |
| Fasting time | | 1.04 | 0.068 | 1.00 | 1.09 |

**Table S9h: Cox proportional hazard analyses of time from registration to death with IPF** with respect to (i) LTL adjusted for age only (amongst all males), (ii) LTL adjusted for age and free testosterone % and (iii) LTL adjusted for age and free testosterone % with additional adjustment for sex hormone level confounders, (iv) LTL adjusted for age and free testosterone concentration and (v) LTL adjusted for age and free testosterone concentration with additional adjustment for sex hormone level confounders. Testing showed no evidence that the proportional hazards assumptions were violated.

| Trait | Sample size | HR | P | 95% CI | |
|---|---|---|---|---|---|
| (i) LTL log adjusted only for age | 694 | 0.93 | 0.098 | 0.85 | 1.01 |
| Age | | 1.04 | $4.9x10^{-4}$ | 1.02 | 1.07 |
| (ii) LTL log adjusted for SHBG | 682 | 0.94 | 0.24 | 0.86 | 1.04 |
| Age | | 1.05 | $4.4x10^{-4}$ | 1.02 | 1.07 |
| SHBG | | 1.002 | 0.46 | 0.996 | 1.008 |
| (iii) LTL log with multiple covariates | 586 | 0.94 | 0.24 | 0.86 | 1.04 |
| Age | | 1.04 | 0.0013 | 1.02 | 1.07 |
| SHBG | | 1.002 | 0.57 | 0.995 | 1.008 |
| Ever smoker | | 1.08 | 0.64 | 0.79 | 1.46 |
| Body mass index | | 0.99 | 0.74 | 0.96 | 1.03 |
| Social deprivation | | 1.00 | 0.83 | 0.96 | 1.03 |
| Fasting time | | 1.04 | 0.096 | 0.99 | 1.08 |

**Table S9i: Cox proportional hazard analyses of time from registration to death with IPF** with respect to (i) LTL adjusted for age only (amongst all males), (ii) LTL adjusted for age and SHBG and (iii) LTL adjusted for age and SHBG with additional adjustment for sex hormone level confounders. Testing showed no evidence that the proportional hazards assumptions were violated.
